# Deep-learning body composition analysis of routine CT robust to scan range, slice order and contrast phase

**DOI:** 10.64898/2026.11.12.26367971

**Authors:** Shouhei Hanaoka, Wataru Gonoi, Shohei Inui, Nobuhiko Akamatsu, Yukihiro Nomura, Tomomi Takenaga, Soichiro Miki, Takeharu Yoshikawa, Kotaro Sugawara, Satoru Taguchi, Kenjiro Kishitani, Haruki Kume, Taketo Kawai, Tohru Nakagawa, Osamu Abe

**Author notes:** **Corresponding author:** Shouhei Hanaoka, Department of Radiology, Graduate School of Medicine, The University of Tokyo, 7-3-1 Hongo, Bunkyo-ku, Tokyo, Japan.

## Abstract

**Purpose:** To develop and validate a fully automated pipeline that measures skeletal muscle and adipose tissue at L1 and L3 on routine CT with varied scan ranges, slice orders and contrast phases, rejects levels outside the scan and converts measures to non-contrast equivalents.

**Materials and methods:** The pipeline (sarco) combines orientation detection, L1/L3 localisation with out-of-range rejection, U-Net segmentation, six-class contrast-phase estimation, phase-specific conversion to non-contrast-equivalent values and, as an auxiliary output, muscle and marrow age estimation. We evaluated it on five single-centre datasets (4986 series): agreement with radiologist-corrected labels using five-fold cross-fitting (2025 series), artificially truncated scans, contrast-enhanced series paired with non-contrast series of the same examination, and the prognostic value of fully automated measures in 2006 patients after curative cancer surgery. TotalSegmentator served as a comparator.

**Results:** On held-out folds, L3 skeletal muscle area (SMA) agreed with the reference (intraclass correlation coefficient 0.983; bias +1.35 cm²). On truncated scans processed with cross-fitted localisation models, the chosen L3 slice was off by half a vertebra or more in 4.7–5.6% of volumes (TotalSegmentator 11.3–14.3%). Four-group phase accuracy was 0.949, exceeding TotalSegmentator in scans shorter than 400 mm (0.952–0.960 vs 0.643–0.668), and conversion reduced the root mean square error of L3 muscle attenuation against the same-examination non-contrast series from 7.96 to 2.81 HU. Combined age estimates had a mean absolute error of 6.5 years. Automatically measured low muscle mass increased the cross-validated Uno’s C of the baseline model by 0.034 (95% CI 0.009–0.053; cancer-specific survival) and 0.031 (0.016–0.048; overall survival), whereas converted L3 muscle attenuation added 0.012 (−0.005 to 0.024) for overall survival.

**Conclusion:** The pipeline provides robust, contrast-phase-aware body composition measures from routine CT with varied scan ranges and reproduces known prognostic associations without manual input.

**Short abstract:** We validated a fully automated pipeline for L1/L3 body composition on routine CT. It handled truncated scans, estimated the contrast phase, converted measures to non-contrast equivalents, agreed closely with radiologist-corrected labels and reproduced prognostic associations in 2006 surgical patients.

## Introduction

Body composition measured on CT, in particular the skeletal muscle area (SMA) and mean skeletal muscle density (MSMD) at the third lumbar vertebra (L3), is associated with outcomes in many cancers and is increasingly used as an opportunistic imaging biomarker [1–3]. In our institution, low muscle mass on preoperative CT was associated with cancer-specific and overall survival after curative surgery for gastrointestinal and urological cancers [4], and low muscle attenuation with overall survival in the same cohort [5]. Manual or semi-automatic measurement is, however, time-consuming.

Several deep-learning tools now segment muscle and adipose tissue on CT, including the widely used open-source TotalSegmentator [6]. Three practical problems remain. First, most tools assume that the target vertebral level is within the scan; studies of automatic L3 selection have often excluded examinations in which L3 was not imaged [7], so the behaviour of these tools on short scans, such as upper-abdominal dynamic CT, is largely unknown. Second, the chosen slice affects the measured area [8], so comparisons between tools are confounded by different slice definitions. Third, iodinated contrast medium increases muscle attenuation by a few to more than 10 Hounsfield units (HU) depending on the phase [9–12], whereas published attenuation thresholds were derived from non-contrast CT [13], and existing corrections were derived for a single phase [10] or apply one phase-independent offset [14]; neither estimates the phase from the image.

We developed sarco, a pipeline that runs as a plug-in of the CIRCUS research platform [15]. It detects the slice order, localises L1 and L3 or reports that they are out of range, segments muscle and vertebral bone, estimates the contrast phase from the image and converts measures to non-contrast (phase 0) equivalents. As auxiliary outputs, it also estimates muscle and marrow age, height and weight; these are summarised in the Online Resource, and the focus of this study is robustness to scan range, slice order and contrast phase. We evaluated (1) its robustness to slice order and truncated scans; (2) its agreement with radiologist-corrected labels, using cross-fitting; (3) phase estimation and phase-0 conversion against same-examination non-contrast series; and (4) whether its fully automated measures reproduce published prognostic associations. TotalSegmentator was used as a comparator where its outputs were comparable.

## Materials and methods

### Ethics and study design

This retrospective single-centre study was approved by the Research Ethics Committee of the Faculty of Medicine of the University of Tokyo (approval number 2561-(30), first approved on 25 May 2009, and, for the prognostic cohort, 2022046NI). The requirement for written informed consent was waived, and patients could opt out. The primary prognostic comparisons were specified in writing on 22 October 2026, before the cross-fitted, fully automated analysis was run, but after exploratory analyses of the same cohort with the reference labels; the study was not pre-registered. Reporting follows the Checklist for Artificial Intelligence in Medical Imaging (CLAIM) 2024 update [16] and, for the prognostic part, the STROBE statement [17] (Online Resource, Tables S9 and S10).

### Datasets

Five datasets of CT examinations performed at The University of Tokyo Hospital were used (Table 1). All images were pseudonymised (patient identifiers replaced by study codes) and converted to NIfTI; the slice thickness equalled the slice interval in all series. Sample sizes were determined by the available data; no power calculation was performed.

- **Dataset main** (2039 patients, one preoperative series each; CT from 2005–2018): patients who underwent curative surgery for gastric, oesophageal, renal or urothelial cancer; 2015 of them were previously reported [4, 5]. L1 and L3 labels of skeletal muscle and vertebral cortical and medullary bone were available (see Reference labels), together with measured height and weight and outcomes.
- **Dataset L** (321 patients, 1675 series, 2022): multiphase abdominal CT acquired with a fixed protocol of non-contrast, early arterial, late arterial, portal venous and equilibrium phases; 203 of these examinations were previously reported [9].
- **Dataset P** (99 patients, 222 series, 2025): pancreatobiliary dynamic CT with non-contrast, venous and equilibrium phases.
- **Dataset HC** (474 patients, 474 series, 2007–2008): whole-body non-contrast CT from health-screening positron emission tomography/CT.
- **Dataset O** (144 patients, 576 series, 2025): other clinical CT with various phases and scan ranges, including head and neck, chest and abdominal examinations.

**Table 1.**
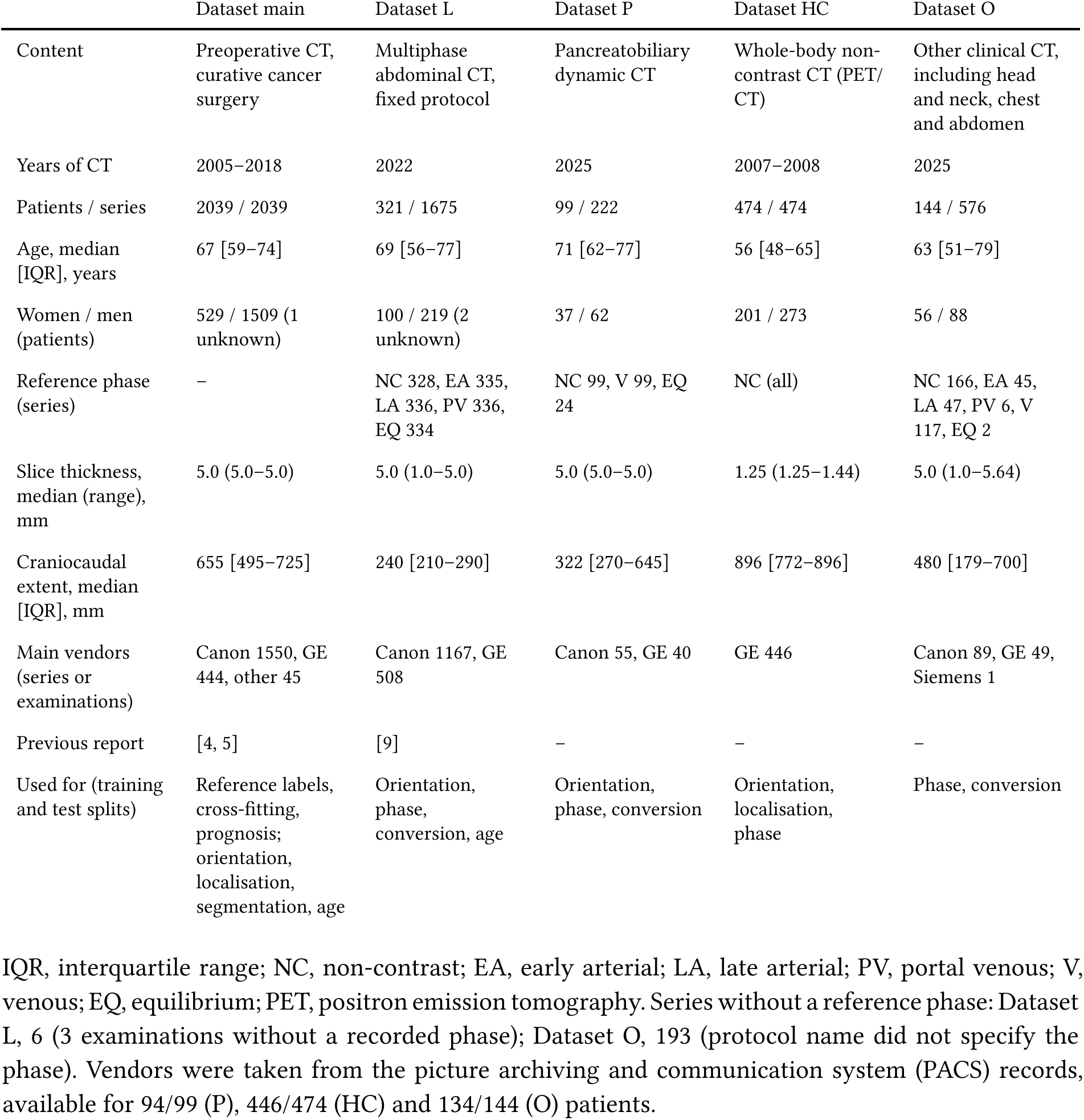
Datasets.

|  | Dataset main | Dataset L | Dataset P | Dataset HC | Dataset O |
|---|---|---|---|---|---|
| Content | Preoperative CT, curative cancer surgery | Multiphase abdominal CT, fixed protocol | Pancreatobiliary dynamic CT | Whole-body non-contrast CT (PET/CT) | Other clinical CT, including head and neck, chest and abdomen |
| Years of CT | 2005–2018 | 2022 | 2025 | 2007–2008 | 2025 |
| Patients / series | 2039 / 2039 | 321 / 1675 | 99 / 222 | 474 / 474 | 144 / 576 |
| Age, median [IQR], years | 67 [59–74] | 69 [56–77] | 71 [62–77] | 56 [48–65] | 63 [51–79] |
| Women / men (patients) | 529 / 1509 (1 unknown) | 100 / 219 (2 unknown) | 37 / 62 | 201 / 273 | 56 / 88 |
| Reference phase (series) | – | NC 328, EA 335, LA 336, PV 336, EQ 334 | NC 99, V 99, EQ 24 | NC (all) | NC 166, EA 45, LA 47, PV 6, V 117, EQ 2 |
| Slice thickness, median (range), mm | 5.0 (5.0–5.0) | 5.0 (1.0–5.0) | 5.0 (5.0–5.0) | 1.25 (1.25–1.44) | 5.0 (1.0–5.64) |
| Craniocaudal extent, median [IQR], mm | 655 [495–725] | 240 [210–290] | 322 [270–645] | 896 [772–896] | 480 [179–700] |
| Main vendors (series or examinations) | Canon 1550, GE 444, other 45 | Canon 1167, GE 508 | Canon 55, GE 40 | GE 446 | Canon 89, GE 49, Siemens 1 |
| Previous report | [4, 5] | [9] | – | – | – |
| Used for (training and test splits) | Reference labels, cross-fitting, prognosis; orientation, localisation, segmentation, age | Orientation, phase, conversion, age | Orientation, phase, conversion | Orientation, localisation, phase | Phase, conversion |

IQR, interquartile range; NC, non-contrast; EA, early arterial; LA, late arterial; PV, portal venous; V, venous; EQ, equilibrium; PET, positron emission tomography. Series without a reference phase: Dataset L, 6 (3 examinations without a recorded phase); Dataset O, 193 (protocol name did not specify the phase). Vendors were taken from the picture archiving and communication system (PACS) records, available for 94/99 (P), 446/474 (HC) and 134/144 (O) patients.

Datasets P, HC and O have not been reported previously. The reference phase was defined by the protocol in Dataset L, was non-contrast in all of Dataset HC, and was taken from the DICOM series description and the protocol name in the radiology information system in Datasets P and O. A few patients appeared in more than one dataset (Online Resource, Fig. S2). Scanners from five vendors were used (Online Resource, Table S2).

### Reference labels

Reference labels in Dataset main were produced as previously described [4, 18]: a first version of the segmentation model was trained on manually painted cases (ITK-SNAP [19]), applied to all cases in CIRCUS, and its output was corrected by radiologists. The reference slice for each level was the slice on which both transverse processes were visible. Because the reference labels originate from an earlier version of the same model family, agreement may favour the proposed pipeline over other tools.

### Pipeline

The pipeline (Fig. 1) processes one CT series as follows; model details are given in Online Resource, Methods S1 and Table S1.

1. **Orientation.** Header orientation information was unreliable: the stored craniocaudal direction of all Dataset L series did not match the image content, and all Dataset HC series were stored feet first. A small convolutional neural network classifies the craniocaudal order from coronal and sagittal mean and maximum intensity projections; the volume is reversed only if the probability of feet first exceeds 0.7, because reversing a correctly ordered volume is the more harmful error.
2. **L1/L3 localisation.** A U-Net [20] applied to projection images outputs, for each craniocaudal position, the likelihood of being L1 or L3. It was trained on randomly cropped scan ranges (120 mm to the whole volume) so that it also learns the absence of a level. A level is reported as out of range when the likelihood peak is below a threshold chosen on validation data, or when it lies within 10 mm of the first or last slice (edge rule).
3. **Segmentation.** A two-dimensional U-Net [20] segments skeletal muscle and vertebral cortical and medullary bone on the selected slice (1 mm pixels, 512 × 512).
4. **Measures.** SMA is the area of muscle pixels between −29 and 150 HU, and MSMD is their mean attenuation; the skeletal muscle index (SMI) is SMA divided by height squared. A visceral region mask (the region enclosed by muscle, excluding muscle and vertebra) is produced. Visceral adipose tissue (VAT) comprises pixels of −150 to −50 HU in the visceral region, and subcutaneous adipose tissue (SAT) pixels of −190 to −30 HU outside the visceral region and muscle; their areas and mean attenuations are computed, as are the proportion of all muscle-label pixels below −30 HU and vertebral bone attenuation.
5. **Contrast phase.** A 2.5-dimensional ResNet18 [21] with convolutional block attention modules [22] (ct25d-regressor) classifies six phases (non-contrast, early arterial, late arterial, portal venous, venous, equilibrium) from the visceral region at L1 and L3; the class probabilities of the available levels are averaged.
6. **Phase-0 conversion.** Each measure is converted to its non-contrast equivalent (*v*_0_) from the value in phase *k* (*v_k_*) with the equation of the estimated phase: a linear equation (*v*_0_ = *a* + *b* × *v_k_*) for attenuation values and proportions, and a power function (*v*_0_ = *c* × *v_k_^p^*) for SMA, VAT and SAT areas, which keeps converted areas positive. Equations were fitted on same-examination pairs of non-contrast and phase-*k* series from Datasets L, O and P; for evaluation, they were refitted on the training and validation pairs only (Online Resource, Table S11).
7. **Age and body size.** Muscle and marrow ages at L1 and L3, height and weight are estimated with the same 2.5-dimensional network (Online Resource, Methods S2 and S4). All analyses in this study used measured height.

**Fig. 1.**
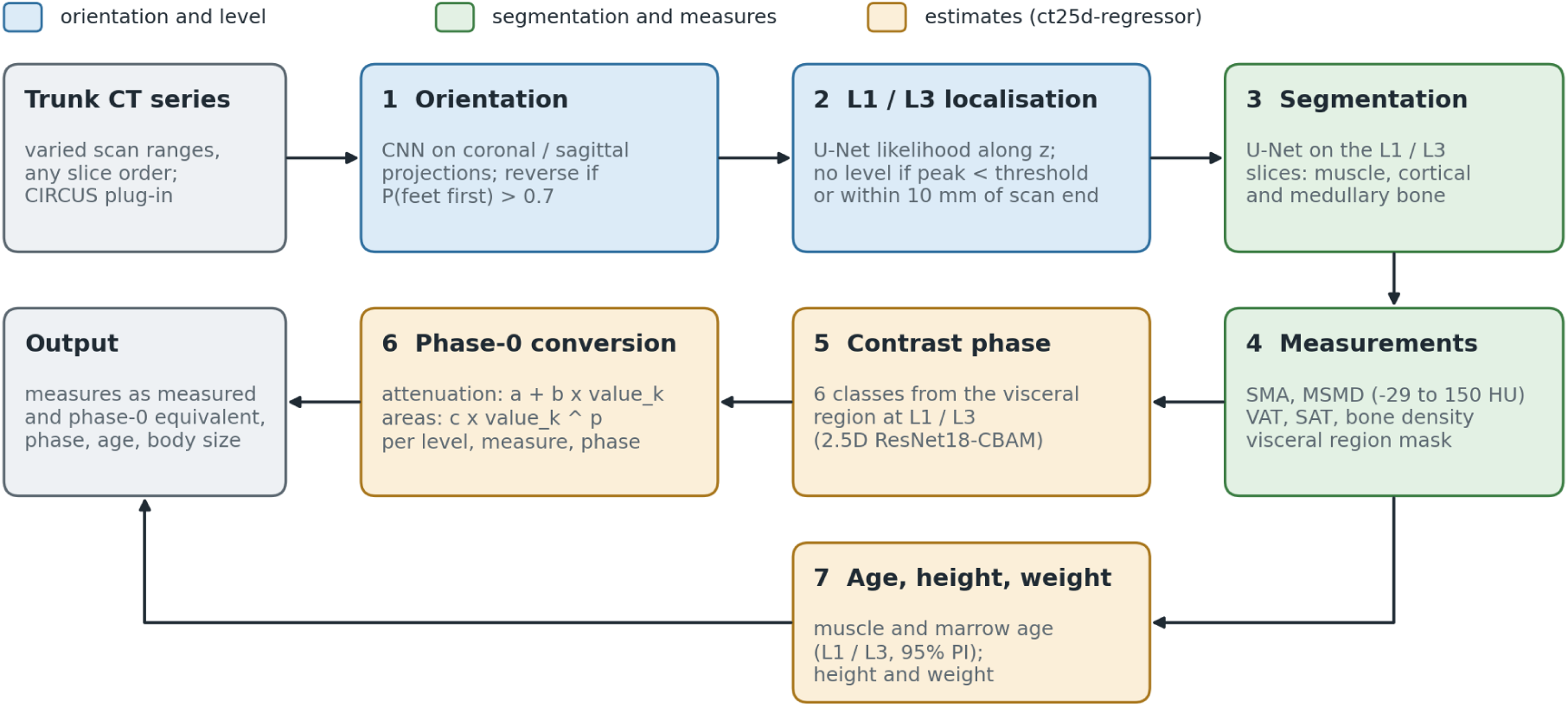
Pipeline of sarco. Each CT series is checked for slice order (1), L1 and L3 are localised or reported as out of range (2), muscle and vertebral bone are segmented on the selected slices (3), and the measures and a visceral region mask are computed (4). The contrast phase is estimated from the visceral region (5) and used to convert measures to non-contrast (phase 0) equivalents (6); muscle and marrow ages, height and weight are also estimated (7). Outputs include the measures, the estimated phase, ages and body size. All results are mapped back to the original slice order.

Results are mapped back to the original slice order. Examples of the output are shown in Fig. 2. Processing one series took a median of 63 s (interquartile range 60–65 s; one graphics processing unit, including reading and writing). All results correspond to the released version, sarco v8.6.0 (version history in Online Resource, Methods S1). The software (sarco and ct25d-regressor) and the trained weights are publicly available (see Data availability); sarco is a research tool and has not been approved as a medical device.

**Fig. 2.**
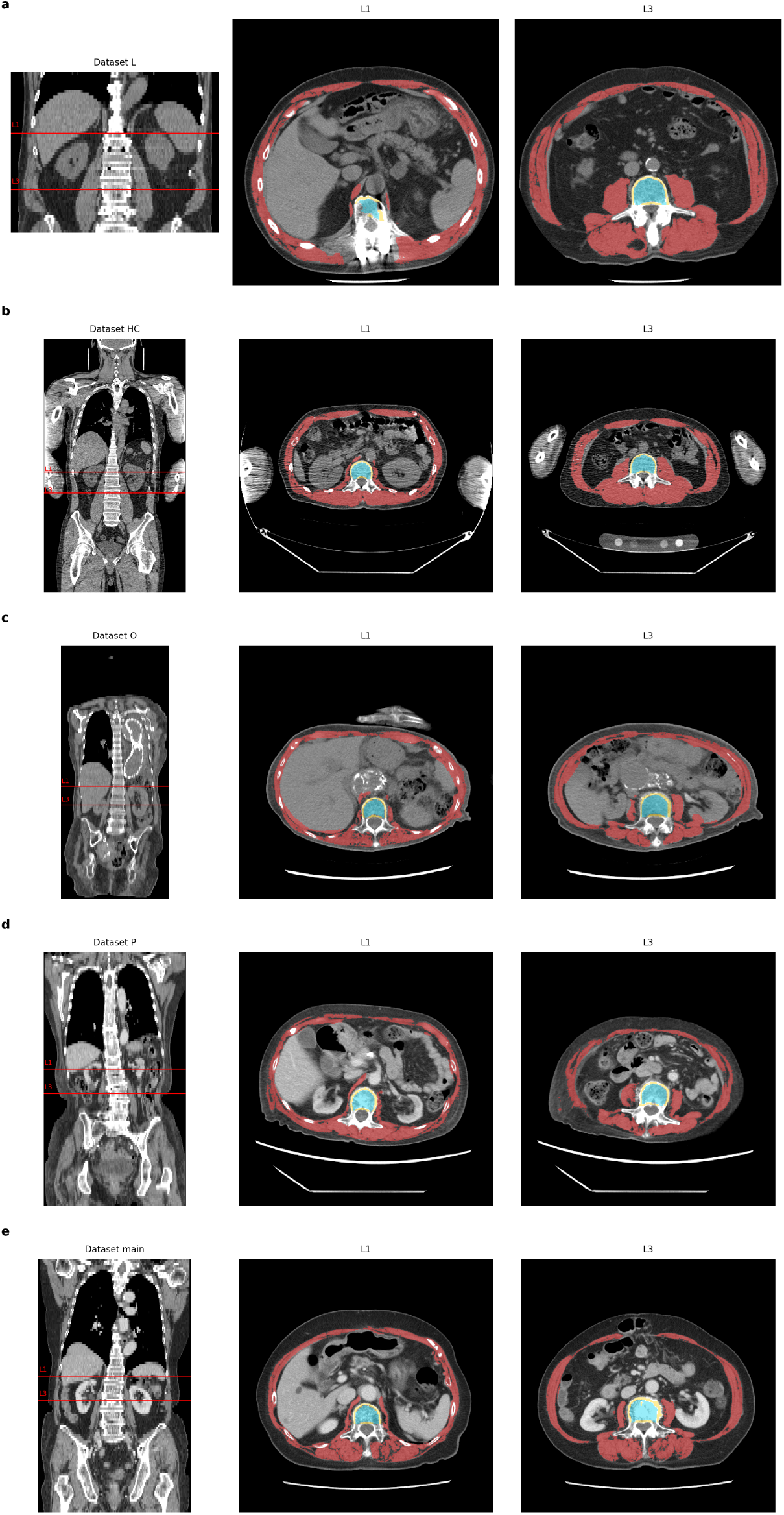
Examples of the output in the five datasets (a, Dataset L; b, Dataset HC; c, Dataset O; d, Dataset P; e, Dataset main): coronal reformation with the detected L1 and L3 levels, and the L1 and L3 slices with skeletal muscle (red), cortical bone (yellow) and medullary bone (cyan).

### Evaluation

#### Orientation and level localisation

The orientation classifier and the localisation models were evaluated on held-out test splits (patient-level 70/15/15 split); the orientation classifier was also evaluated on cropped scans and compared with an earlier version trained only on abdominal CT (Online Resource, Methods S1). For truncation robustness, the test-split series of Dataset main (5-mm slices) were cut to 200 mm so that the target level was (a) in the whole, uncut scan, (b) 80–120 mm from the start of the cut, (c) 10–50 mm inside its first or last slice, (d) 5–30 mm beyond it or (e) 30–100 mm beyond it (274–303 volumes per scenario and level). For each in-range scenario we recorded whether the level was found and whether the chosen slice was off by at least half a vertebra (16 mm, half the median vertebral height); for out-of-range scenarios, any reported level was a false detection. Each volume was processed with the cross-fitted localisation models (see below) that had not been trained on that patient; results with the deployed models and without the edge rule are given in Online Resource, Table S12. All datasets were also processed as they were to count series in which L1 and L3 were found.

#### Cross-fitting and agreement

Because the deployed segmentation and localisation models had been trained on all of Dataset main, both were retrained five times from random initialisation, each time leaving out one fold (folds stratified by cancer type and cancer-specific death), with otherwise unchanged settings. Each series was then processed by the models that had not seen it. Fourteen series with inconsistent reference labels were excluded, leaving 2025. We evaluated (a) segmentation on the reference slice (Dice, SMA, MSMD) and (b) the whole pipeline with its own slice selection (slice distance, SMA, SMI, MSMD, and low muscle mass by the criteria of Fujiwara et al. [23]: L3 SMI < 36.2 cm²/m² for men and < 29.6 cm²/m² for women). Agreement was summarised by the intraclass correlation coefficient (ICC) for absolute agreement (ICC(A,1)) and for consistency (ICC(C,1)) [24, 25] and by Bland–Altman bias and 95% limits of agreement (LoA). The effect of level selection was assessed by the distance between the chosen and reference slices (Online Resource, Table S5).

#### Comparison with TotalSegmentator

TotalSegmentator 2.18.0 [6, 26] (TotalSegmentator Non-Commercial License) was applied to the same series for comparison only: the L3 vertebra from the `total` task, skeletal muscle from the `tissue_types` task and the phase from `totalseg_get_phase`, which predicts the time after injection and maps it to four classes without an equilibrium class. Its L3 slice was the craniocaudal middle of its L3 mask. Phase accuracy was compared in four groups (non-contrast, arterial, portal venous/venous, equilibrium) on the test splits of Datasets L, HC, O and P. Details are given in Online Resource, Methods S5.

#### Phase-0 conversion

In the test splits, contrast-enhanced series were paired with the non-contrast series of the same examination (L3 MSMD, 204 pairs; L1, 237 pairs). The contrast-series value was compared with the non-contrast value as measured, after conversion with the estimated phase and after conversion with the reference phase. Because the thresholds for low muscle attenuation used in the prognostic cohort [5] originate from non-contrast CT of healthy young adults [13], we also counted how classification changed after conversion (Online Resource, Table S6). Phase-specific differences as measured were also summarised in all patients of Datasets L and P, using the earliest examination per patient (Online Resource, Table S15).

#### Prognosis

In Dataset main, all predictors were taken from the cross-fitted pipeline (fully automated; missing levels treated as missing), with SMI computed from measured height. Because the Fujiwara thresholds were derived mainly from contrast-enhanced CT, SMI was computed from areas as measured; a sensitivity analysis used phase-0 areas. Endpoints were cancer-specific survival (CSS) and overall survival (OS) from surgery; follow-up ended in December 2022 (median 61 months; interquartile range 34–79) [4]. Two primary analyses were defined: (1) the addition of Fujiwara low muscle mass to the published baseline model (age ≥ 70 years, sex, urological cancer, stage IV) for CSS and OS (A1 vs A0); and (2) the addition of phase-0 L3 MSMD (per 10 HU) to the baseline model with body mass index (BMI) for OS (C4 vs C0). Cox proportional hazards models were fitted, and proportional hazards were checked with Schoenfeld residuals. Discrimination was measured by Uno’s C at 60 months [27], both apparent and cross-validated (10-fold, 20 repeats, identical folds for all models). The cross-validated difference in C (ΔC) was the primary measure of added value; its 95% CI was obtained from 500 patient-level bootstrap samples, in each of which the whole cross-validation (10 folds, 5 repeats) was repeated. Secondary analyses, including muscle and marrow age gaps, are described in Online Resource, Methods S3.

#### Statistics

Values are mean ± standard deviation (SD) unless stated otherwise. Analyses used Python 3.12, PyTorch 2.11 and statsmodels 0.15.0; Uno’s C and ICCs were computed with our own implementations, which were checked against reference implementations. Two-sided P < 0.05 was considered significant; secondary analyses were not adjusted for multiplicity and are exploratory.

#### Use of generative artificial intelligence

Claude and Claude Code (Anthropic, San Francisco, CA, USA) were used to assist in writing analysis code and drafting the manuscript. The authors reviewed all outputs and take full responsibility for the content.

## Results

### Datasets and level detection

Table 1 summarises the datasets. With the deployed pipeline, both L1 and L3 were found in 2024 of 2039 series of Dataset main, 1383 of 1675 of Dataset L, 220 of 222 of Dataset P, 463 of 474 of Dataset HC (the localisation models were partly trained on visually confirmed Dataset HC series) and 362 of 576 of Dataset O. Missed levels were concentrated in short scans: in Dataset L, 143 of 258 series covering less than 200 mm yielded L1 only (upper-abdominal dynamic CT), and in Dataset O, 149 of 160 series covering less than 200 mm yielded neither level (Online Resource, Fig. S1).

### Orientation

On the test splits, no correctly ordered series was reversed in any dataset. Missed feet-first series were rare (0.8% for Dataset L cropped to 200 mm, 0.3% for Dataset main cropped to 300 mm and 1 of 71 whole-body series of Dataset HC; none otherwise). A classifier trained only on abdominal CT reversed 52% of correctly ordered whole-body series, and the broader training raised the number of Dataset HC series with both levels found from 136 to 439 of 474 (with the earlier localisation models; 463 with the final models, see above).

### Truncated scans

When the level was in view, the cross-fitted models found it in 97.7–100% of volumes and chose the same slice as the reference in 73.2–74.3% (L1) and 71.0–72.4% (L3), similar to whole scans in the agreement analysis (Fig. 3). The chosen L3 slice was off by half a vertebra or more in 5.6%, 5.5% and 4.7% of whole, mid-range and near-end volumes (L1: 4.3%, 4.9% and 3.7%), compared with 11.3%, 11.7% and 14.3% for TotalSegmentator (L1: 7.3%, 4.2% and 10.0%); because TotalSegmentator’s rate was similar on whole, uncut scans, it partly reflects its different slice definition rather than truncation. When the level was just outside the scan, false detections occurred in 7.1% (L1) and 13.2% (L3) of volumes with sarco and in 10.4% and 13.6% with TotalSegmentator; without the edge rule, they rose to 33.0% and 26.1% for sarco. When the level was far outside, false detections were 0–0.7% for both tools. The deployed models, partly trained on these patients, performed much better (L3 off by half a vertebra or more in 0–0.7%; Online Resource, Table S12).

**Fig. 3.**
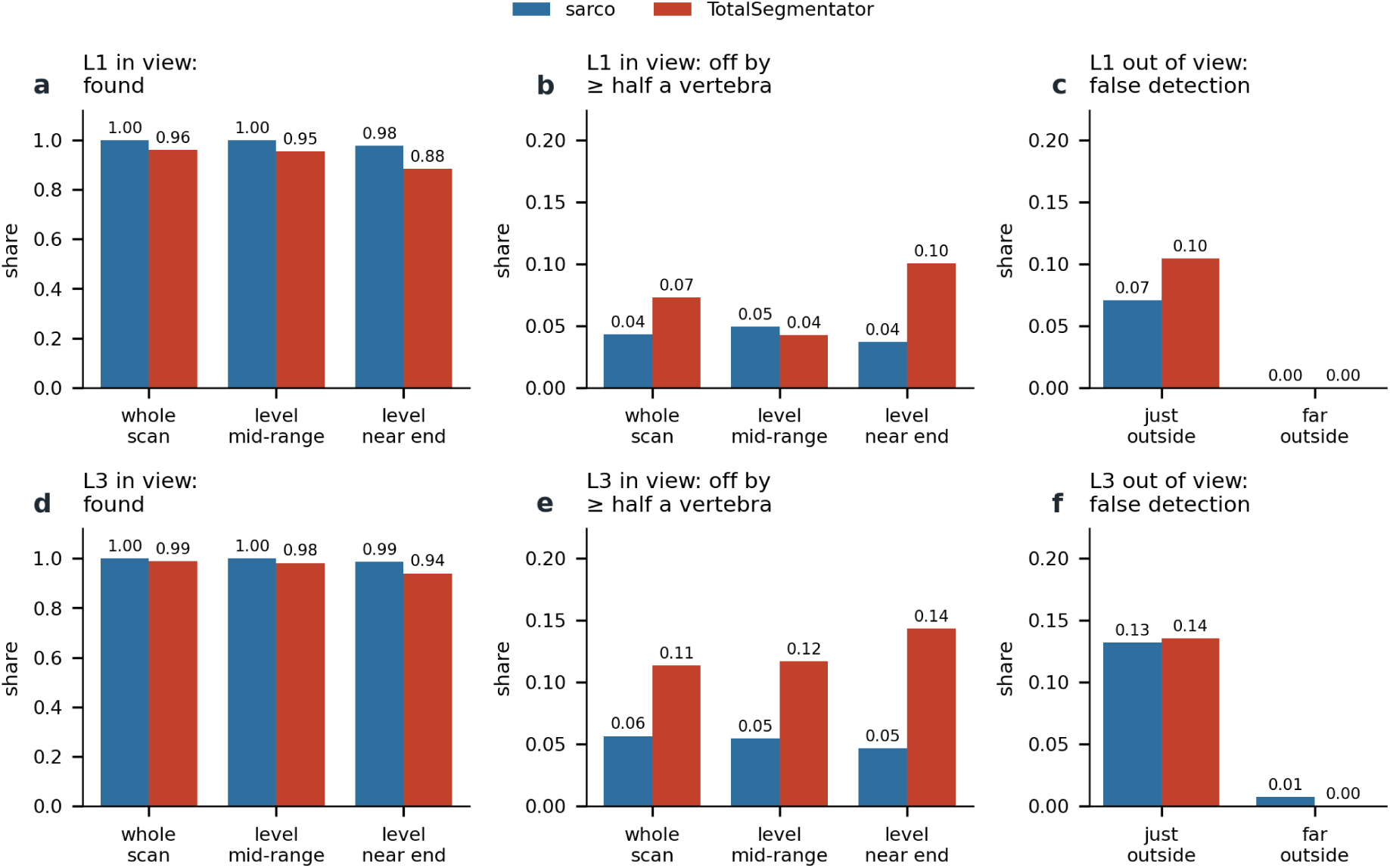
Truncated test scans. Proportion of volumes in which the level was found (a, d) and in which the chosen slice was off by at least half a vertebra (b, e) when L1 (a–c) or L3 (d–f) was in view, and proportion of false detections when the level was just (5–30 mm) or far (30–100 mm) outside the scan (c, f), for sarco (cross-fitted localisation models, with the edge rule) and TotalSegmentator.

### Agreement with the reference labels

On the reference slice, the cross-fitted U-Net achieved a muscle Dice of 0.944 ± 0.028 at both levels (0.964 ± 0.022 at L3 after the −29 to 150 HU threshold) (Table 2). L3 SMA agreed closely (ICC(A,1) 0.988; bias +1.15 cm²; LoA −6.27 to 8.57 cm²), as did MSMD (ICC(A,1) 0.997; bias −0.18 HU). With automatic slice selection, the L3 slice was identical to the reference in 72.8% of series, within ±1 slice in a further 22.3% and within 5 mm in 95.3% of the 2007 series in which L3 was found. Agreement of the whole pipeline remained high for SMA (L3 ICC(A,1) 0.983; bias +1.35 cm²; LoA −7.44 to 10.14 cm²), SMI (ICC(A,1) 0.972) and MSMD (ICC(A,1) 0.991), with similar results at L1 (Table 2; in-sample values of the deployed models in Online Resource, Table S3). Classification of low muscle mass agreed in 95.5% of patients (κ = 0.838); the pipeline classified fewer patients as having low muscle mass (low by the reference only, 66; by the pipeline only, 24).

**Table 2.**
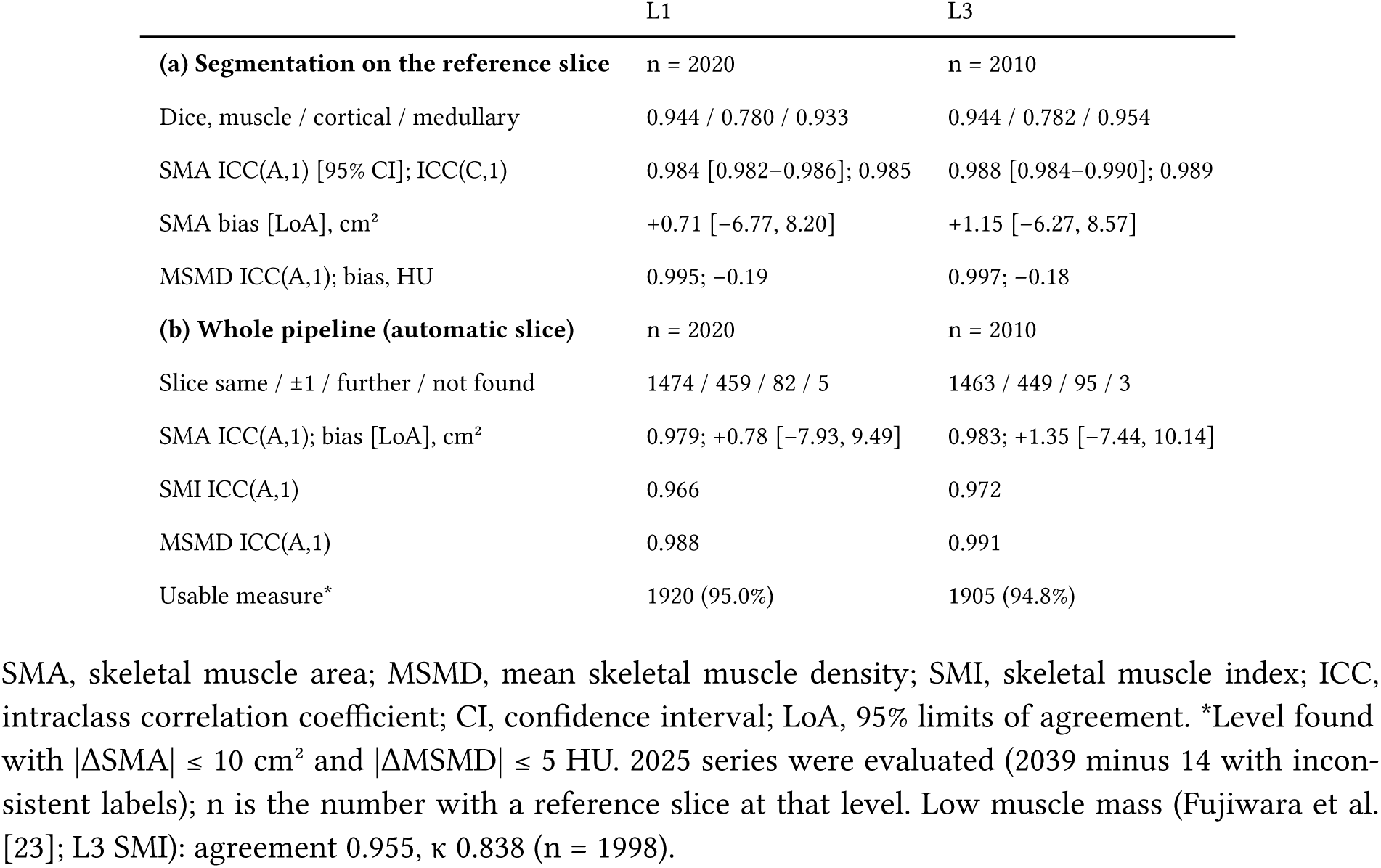
Agreement with the reference labels in Dataset main (cross-fitted models; 2025 series)

|  | L1 | L3 |
|---|---|---|
| **(a) Segmentation on the reference slice** | n = 2020 | n = 2010 |
| Dice, muscle / cortical / medullary | 0.944 / 0.780 / 0.933 | 0.944 / 0.782 / 0.954 |
| SMA ICC(A,1) [95% CI]; ICC(C,1) | 0.984 [0.982−0.986]; 0.985 | 0.988 [0.984−0.990]; 0.989 |
| SMA bias [LoA], cm² | +0.71 [−6.77, 8.20] | +1.15 [−6.27, 8.57] |
| MSMD ICC(A,1); bias, HU | 0.995; −0.19 | 0.997; −0.18 |
| **(b) Whole pipeline (automatic slice)** | n = 2020 | n = 2010 |
| Slice same / ±1 / further / not found | 1474 / 459 / 82 / 5 | 1463 / 449 / 95 / 3 |
| SMA ICC(A,1); bias [LoA], cm² | 0.979; +0.78 [−7.93, 9.49] | 0.983; +1.35 [−7.44, 10.14] |
| SMI ICC(A,1) | 0.966 | 0.972 |
| MSMD ICC(A,1) | 0.988 | 0.991 |
| Usable measure<sup>*</sup> | 1920 (95.0%) | 1905 (94.8%) |

SMA, skeletal muscle area; MSMD, mean skeletal muscle density; SMI, skeletal muscle index; ICC, intraclass correlation coefficient; CI, confidence interval; LoA, 95% limits of agreement. <sup>\*</sup>Level found with \|ΔSMA\| ≤ 10 cm² and \|ΔMSMD\| ≤ 5 HU. 2025 series were evaluated (2039 minus 14 with inconsistent labels); n is the number with a reference slice at that level. Low muscle mass (Fujiwara et al. [23]; L3 SMI): agreement 0.955, κ 0.838 (n = 1998).

A ±1-slice shift barely changed the measures (SD of the L3 SMA difference 3.64 cm² for the same slice and 5.17 cm² for ±1 slice), whereas selecting another vertebra (4.3% of L3 series) widened the SD to 9.88 cm² (Online Resource, Table S5). A usable measure (level found with |ΔSMA| ≤ 10 cm² and |ΔMSMD| ≤ 5 HU, defined in advance) was obtained in 95.0% (L1) and 94.8% (L3) of series; failures were due to missed levels (5 and 3), another vertebra (31 and 36) and segmentation or value errors (64 and 66) (Fig. 4).

**Fig. 4.**
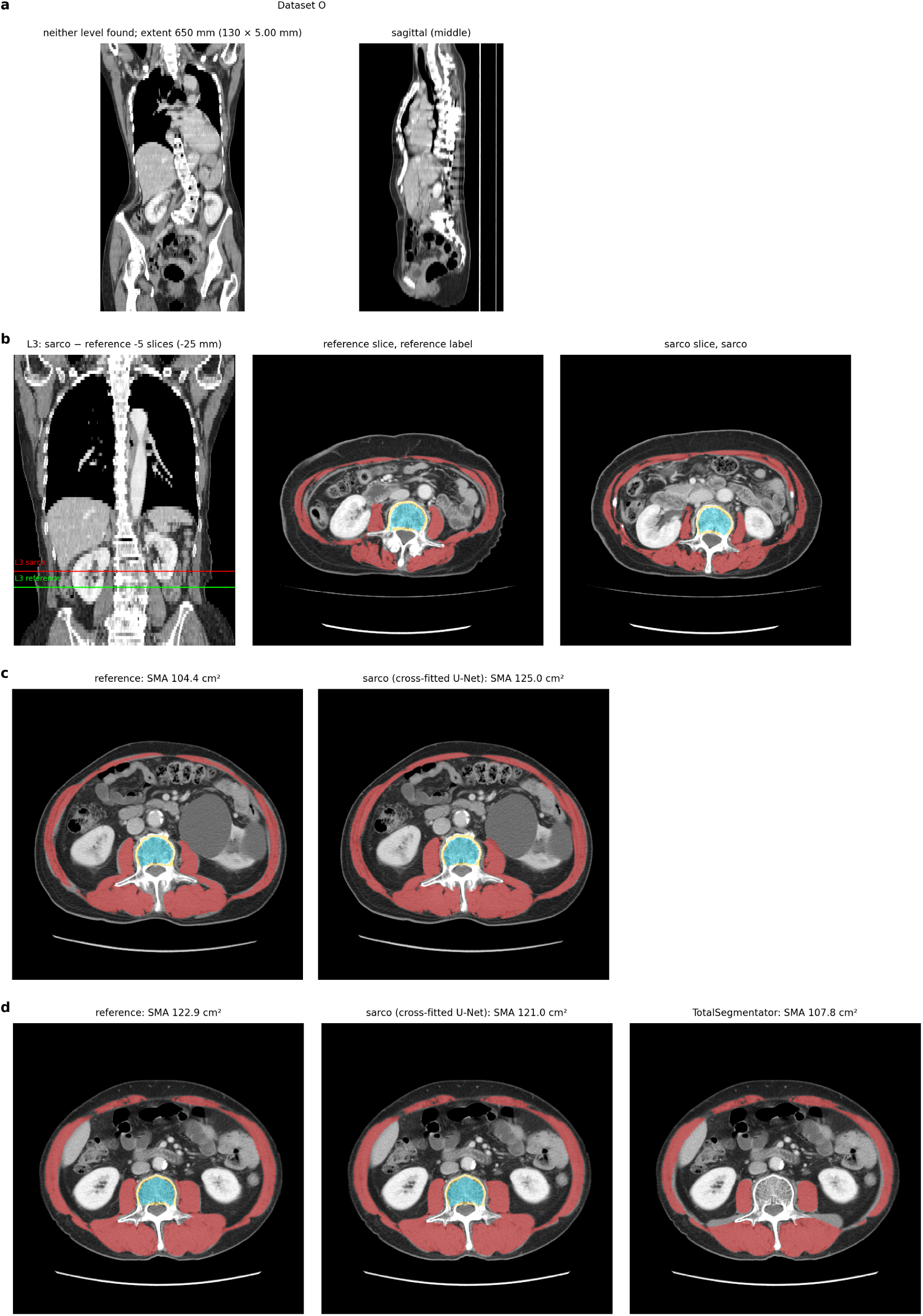
Examples of failure: (a) neither level found (Dataset O), (b) slice selection off the reference, (c) SMA difference above 10 cm² and (d) disagreement with TotalSegmentator.

TotalSegmentator underestimated L3 SMA on the reference slice by 10.7 cm² (9.9% of the reference mean; ICC(A,1) 0.878; ICC(C,1) 0.964), and the underestimation increased with muscle area (Online Resource, Methods S5, Table S13 and Fig. S3).

### Contrast phase and phase-0 conversion

The four-group phase accuracy of sarco was 0.949 (Table 3, Fig. 5). Non-contrast series were separated from contrast-enhanced series almost perfectly (binary accuracy 0.998; accuracy for the non-contrast class 0.993), and venous-phase series were often classified as portal venous or equilibrium. In the 409 series assessed by both methods, sarco was more accurate than TotalSegmentator in series covering less than 200 mm (0.952 vs 0.643) and 200–400 mm (0.960 vs 0.668), whereas TotalSegmentator was more accurate in series covering 400 mm or more (0.982 vs 0.935) (Fig. 5c; Online Resource, Table S4).

**Fig. 5.**
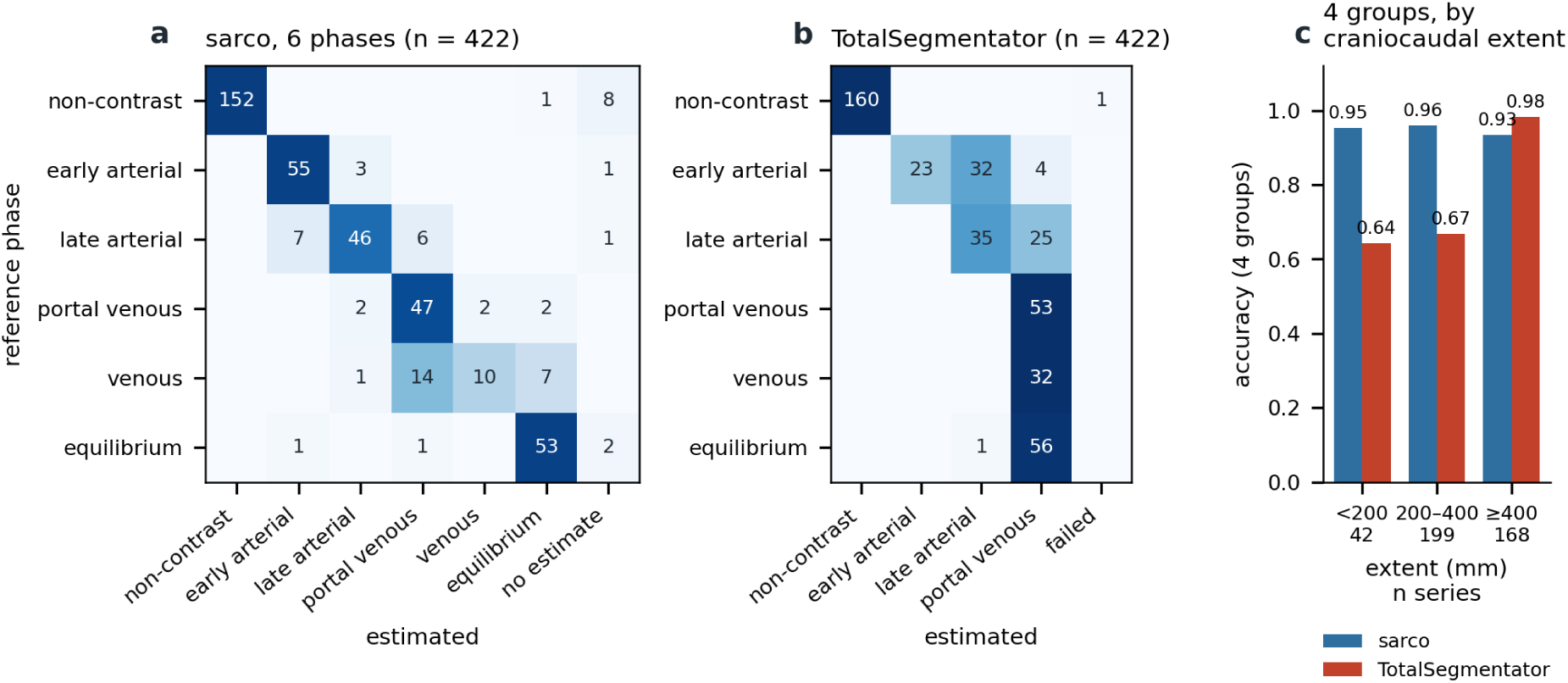
Contrast-phase estimation on the test splits (n = 422 series with a reference phase; 13 series were not assessed by both methods: 12 without a sarco estimate and 1 TotalSegmentator failure). Confusion matrices of sarco (six phases; "no estimate", no phase returned) (a) and TotalSegmentator (four phases; no equilibrium output; "failed", no output) (b) against the reference phase, and four-group accuracy by craniocaudal extent in the 409 series assessed by both methods (c).

**Table 3.** Contrast-phase estimation and phase-0 conversion (test splits)

|  | sarco |  |  |
|---|---|---|---|
| Four-group accuracy (n = 409) | 0.949 |  |  |
| Non-contrast (n = 152)† | 0.993 |  |  |
| Arterial (n = 117) | 0.949 |  |  |
| Portal venous / venous (n = 85) | 0.859 |  |  |
| Equilibrium (n = 55) | 0.964 |  |  |
| Extent < 200 / 200–400 / ≥ 400 mm | 0.952 / 0.960 / 0.935 |  |  |
| Six-phase accuracy, Dataset L / HC / P / O | 0.930 / 1.000 / 0.794 / 0.542 |  |  |
| Contrast − same-examination non-contrast | As measured | Phase 0, estimated phase | Phase 0, reference phase |
| L3 MSMD (204 pairs): bias / SD / RMSE, HU | +6.95 / 3.90 / 7.96 | +0.55 / 2.76 / 2.81 | +0.54 / 2.72 / 2.77 |
| L3 MSMD ICC | 0.688 | 0.940 | 0.942 |
| L1 MSMD (237 pairs): bias / SD / RMSE, HU | +8.06 / 3.82 / 8.92 | +0.71 / 2.69 / 2.77 | +0.68 / 2.65 / 2.73 |

†Class-wise accuracy (proportion of series of each reference group classified correctly) in the 409 series assessed by both methods; non-contrast versus contrast-enhanced binary accuracy, 0.998. Comparison with TotalSegmentator in Online Resource, Table S4. MSMD, mean skeletal muscle density; ICC, intraclass correlation coefficient; SD, standard deviation of the difference; RMSE, root mean square error. Conversion equations were fitted on the training and validation pairs.

Against the non-contrast series of the same examination, contrast-enhanced L3 MSMD was 6.95 HU higher as measured (root mean square error [RMSE] 7.96 HU; ICC 0.688) (Fig. 6). After conversion with the estimated phase, the bias was +0.55 HU and the RMSE 2.81 HU (ICC 0.940), close to conversion with the reference phase (RMSE 2.77 HU). Results at L1 were similar (RMSE 8.92 → 2.77 HU). The estimated phase was wrong in 34 of 204 L3 pairs, but never as non-contrast, and the RMSE was only slightly larger in these pairs (3.02 vs 2.76 HU). For areas, conversion reduced the median absolute error of L3 VAT in phases 2–5 from 21–32% to 7–11%; SMA and SAT changed by 6% or less with contrast, and conversion did not reduce their error (Online Resource, Table S16). Applying the published thresholds for low muscle attenuation to the phase-0 values reclassified a large proportion of the prognostic cohort as low (22.0% as measured vs 56.6% after conversion; Online Resource, Table S6).

**Fig. 6.**
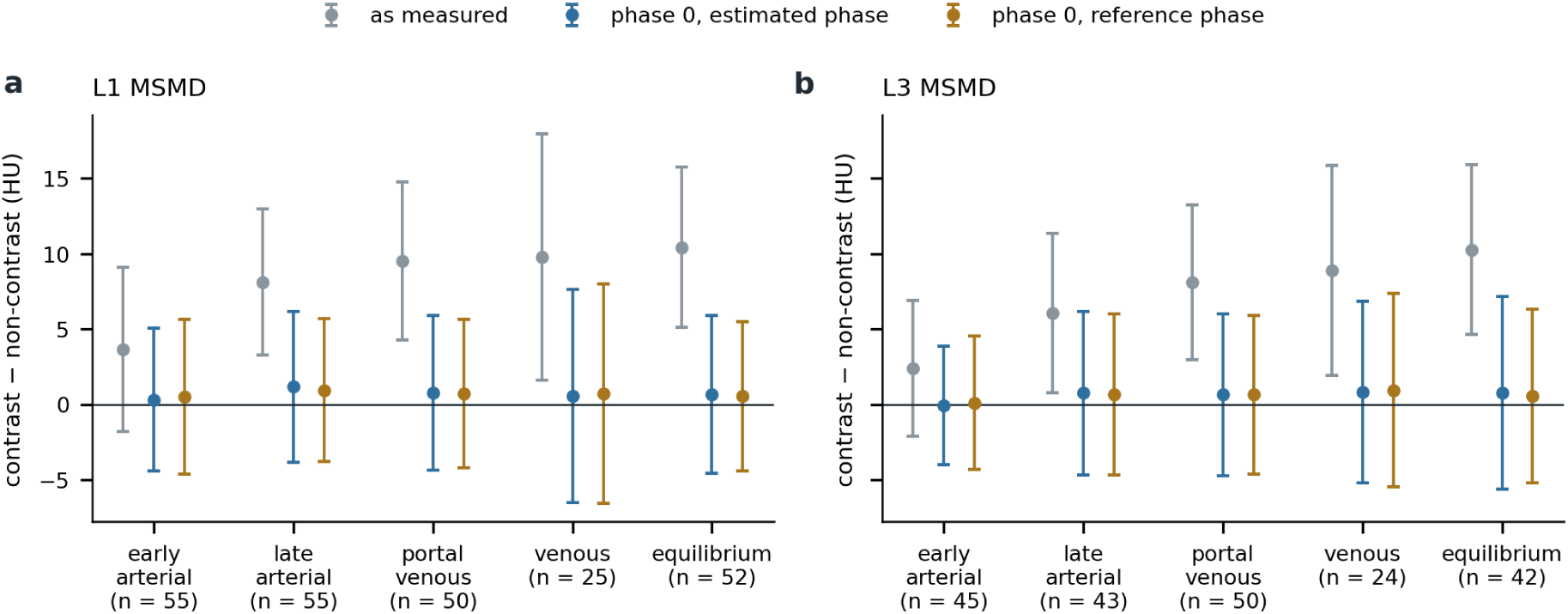
Difference in MSMD between contrast-enhanced series and the non-contrast series of the same examination at L1 (a) and L3 (b), as measured and after conversion to phase-0 equivalents with the estimated or reference phase. Points, mean difference; bars, 95% limits of agreement.

### Prognostic validation

The fully automated analysis included 2006 patients with complete data (302 cancer-specific deaths; 448 deaths) (Table 4). Adding automatically measured low muscle mass to the baseline model increased the cross-validated C from 0.648 to 0.682 for CSS (ΔC +0.034 [95% CI 0.009–0.053]) and from 0.665 to 0.696 for OS (ΔC +0.031 [0.016–0.048]) (hazard ratio [HR] 1.88 [1.43–2.47] and 2.32 [1.88–2.86], respectively). Adding phase-0 L3 MSMD to the baseline model with BMI increased the cross-validated C for OS from 0.700 to 0.712 (ΔC +0.012 [−0.005 to 0.024]; HR per 10 HU 0.61 [0.53–0.70]); for CSS, the gain was +0.005 (HR 0.70 [0.59–0.83]). These results were close to those obtained with the reference labels (apparent C within 0.01 for most models) and with phase-0 areas (Online Resource, Table S14). No covariate showed evidence of non-proportional hazards (all P > 0.14).

**Table 4.** Prognostic analyses with fully automated measures (Dataset main, n = 2006)

|  | CSS (302 events) | OS (448 events) |
|---|---|---|
| A0 baseline: cross-validated C | 0.648 | 0.665 |
| A1 = A0 + low muscle mass: cross-validated C | 0.682 | 0.696 |
| **ΔC A1 – A0 (primary) [95% CI]** | **+0.034 [0.009, 0.053]** | **+0.031 [0.016, 0.048]** |
| HR, low muscle mass [95% CI] | 1.88 [1.43−2.47] | 2.32 [1.88−2.86] |
| C0 = A0 + BMI: cross-validated C | 0.682 | 0.700 |
| C4 = C0 + phase-0 L3 MSMD: cross-validated C | 0.687 | 0.712 |
| **ΔC C4 – C0 (primary for OS) [95% CI]** | +0.005 [−0.016, 0.021] | **+0.012 [−0.005, 0.024]** |
| HR, phase-0 L3 MSMD per 10 HU [95% CI] | 0.70 [0.59−0.83] | 0.61 [0.53−0.70] |
| Published (reference labels), apparent C, baseline → low muscle mass [4]† | 0.63 → 0.67 | 0.66 → 0.70 |

CSS, cancer-specific survival; OS, overall survival; HR, hazard ratio; BMI, body mass index. Uno's C at 60 months; cross-validation 10-fold × 20 repeats; CIs of the cross-validated ΔC from 500 bootstrap samples. Baseline: age ≥ 70 years, sex, urological cancer, stage IV. †Time point of C not reported. Bootstrap CIs of the apparent ΔC and hazard ratios of all models are given in Online Resource, Tables S7 and S8.

### Muscle and marrow age

The combined muscle and marrow age estimate had a mean absolute error of 6.53 years on the pooled test splits of Datasets L and main (n = 554; r = 0.76), and age gaps did not add to the prognostic models (Online Resource, Methods S4 and Fig. S4).

## Discussion

Evaluated out of sample by cross-fitting, the pipeline agreed closely with radiologist-corrected labels and reproduced the prognostic associations previously reported in the same cohort. It handled truncated scans with an accuracy similar to that on whole scans, estimated the contrast phase in short examinations, and converted contrast-enhanced attenuation to non-contrast equivalents with an error below 3 HU.

Automated L3 tools have mostly been evaluated on examinations that include L3; examinations without L3 are often excluded [7], and anatomical variants reduce the success of L3 selection [28]. Vertebral number variants also make the identity of L3 itself ambiguous, and the L3 transverse process is not a landmark independent of the segmentation pattern [29, 30]; our aim was therefore a reproducible measurement position rather than unambiguous vertebral naming. In routine practice many examinations are short, and a tool that always returns a level will silently measure another vertebra. Training the localiser on randomly truncated volumes kept the chosen L3 slice within half a vertebra in about 95% of in-range truncated volumes, as on whole scans, whereas TotalSegmentator, which was not designed for this task and defines the slice differently, was half a vertebra or more away in 11– 14%; at L1 the two tools were closer. The edge rule reduced false detections just outside the scan from 26–33% to 7–13%, a rate similar to TotalSegmentator, so measures from levels close to the scan end still require caution. The much better results of the deployed models show how strongly in-sample evaluation can overstate robustness. To our knowledge, a systematic evaluation of L3 selection on truncated scans has not been reported.

In our data, a one-slice shift had little effect on the measures whereas selecting another vertebra did, consistent with the small differences reported between slices within L3 [8, 31]. TotalSegmentator’s SMA was systematically smaller, as in a multi-tool comparison [31]; its absolute values, and thus thresholds such as that of Fujiwara et al. [23], are not interchangeable with values from corrected manual segmentation, although our reference labels may favour our pipeline.

Contrast enhancement raised L3 muscle attenuation by about 7 HU on average, similar to the portal venous effect of +7.8 HU reported for Dataset L [9]. Existing corrections use a single-phase regression [10] or a phase-independent offset [14], whereas the effect of contrast differs between phases, so that fixed thresholds reclassify patients depending on the phase [32]. Estimating the phase from the image is therefore a prerequisite for phase-specific conversion. The sarco pipeline uses only the visceral region at L1 and L3, so it works in short dynamic examinations, where TotalSegmentator, which infers the time after injection from structures from the head to the pelvis, was less accurate. Its weakest class was the venous phase, but because phase estimation serves to select the conversion, confusion between adjacent late phases had little effect on converted attenuation (RMSE 2.81 vs 2.77 HU with the reference phase). For areas, previous corrections used proportional equations for muscle and fat areas [10, 33] or linear regression [34], and a study of healthy donors advised against mixing contrastenhanced and non-contrast scans within a study because the contrast-related difference in visceral fat area grows with the amount of fat [35]; a power function kept converted areas positive and reduced the error for small VAT.

Phase-0 conversion matters for fixed thresholds. The muscle attenuation thresholds used in our cohort [5] are the mean minus two standard deviations of healthy young adults on non-contrast CT [13], and their authors cautioned against applying them to contrast-enhanced CT. Applied to the phase-0 values, they classified more than half of our older patients with cancer as low and lost much of their prognostic separation, whereas applied to the mostly portal venous values as measured, they reproduced the published prevalence (22.0% vs 21.0% [5]). This suggests that such thresholds need validation in the target population, not that the conversion fails. In our primary analysis, continuous phase-0 MSMD remained associated with OS in a model with BMI, but its cross-validated gain was small and uncertain (ΔC +0.012; 95% CI −0.005 to 0.024).

This study has limitations. First, it is a single-centre study without external validation, and the scanners were mainly from two vendors. Second, the reference labels were produced by correcting the output of an earlier version of the same model family, which may favour our pipeline over TotalSegmentator; cross-fitting removes memorisation of individual cases but not this labelling style, and no independent manual reference was created; inter-reader variability of the reference labels was not assessed. Third, the reference phases in Datasets P and O were derived from series descriptions and protocol names without image-based verification, which may have introduced label noise, particularly for the venous phase. Fourth, the deployed localisation models had been initialised from a model trained on all of Dataset main, so their results on Dataset main are not independent estimates. Fifth, SMI computed from estimated height was not evaluated. Finally, the conversion equations were derived from a limited number of paired examinations, and converted areas were uncertain when VAT was small (median overestimation 14–22% in the lowest quartile at L3 in phases 3–5; Online Resource, Table S16).

## Conclusion

A fully automated pipeline measured L1/L3 body composition on routine CT with varied scan ranges, with explicit rejection of out-of-range levels, estimated the contrast phase and converted attenuation to non-contrast equivalents, agreed closely with radiologist-corrected labels and reproduced known prognostic associations without manual input.

## Acknowledgements

We acknowledge Dr. Jakob Wasserthal, Department of Radiology, University Hospital Basel, and the University Hospital Basel as the source of the TotalSegmentator software, which was used under the TotalSegmentator Non-Commercial License for comparison only. The Department of Computational Diagnostic Radiology and Preventive Medicine, The University of Tokyo Hospital, is sponsored by HIMEDIC Inc. and Siemens Healthcare K.K.

## Ethics approval

This study was approved by the Research Ethics Committee of the Faculty of Medicine of the University of Tokyo (2561-(30); 2022046NI) and conducted in accordance with the Declaration of Helsinki.

## Consent to participate

The requirement for written informed consent was waived because of the retrospective design; patients could opt out.

## Consent for publication

Not applicable; all images are pseudonymised and contain no identifying information.

## Author contributions

Conceptualization: S.H., W.G., S.T., K.S. Methodology: S.H., W.G., S.T., K.S. Software: S.H., Y.N., T.T., S.M. Validation: W.G., S.I., N.A., Y.N., T.T., S.M., T.Y., K.S., S.T., K.K., H.K., T.K., T.N., O.A. Formal analysis: S.H., W.G. Resources: N.A., Y.N., T.T., S.M. Data curation: N.A., Y.N., T.T., S.M. Writing – original draft: S.H. Writing – review and editing: W.G., S.I., N.A., Y.N., T.T., S.M., T.Y., K.S., S.T., K.K., H.K., T.K., T.N., O.A. Visualization: S.H. Supervision: H.K., O.A. Funding acquisition: S.H., W.G. All authors read and approved the final manuscript.

## Data availability

The CT data cannot be shared because of patient privacy. The source code and trained model weights (sarco v8.6.0) are available at https://github.com/utrad-ical/sarco and archived at Zenodo (https://doi.org/10.5281/zenodo.24158133) under the Creative Commons Attribution-Non-Commercial 4.0 International licence. The estimation library ct25d-regressor is available at https://github.com/hanaokalog/ct25d-regressor (commit 0f3b7b1; MIT licence).

## Online Resource

Online Resource 1 (one PDF file) contains:

- Methods S1 (model details; Table S1; orientation evaluation), S2 (height and weight estimation), S3 (secondary and sensitivity analyses), S4 (muscle and marrow age) and S5 (comparison with TotalSegmentator).
- Fig. S1 Series by slice thickness and craniocaudal extent, and whether L1/L3 were found.
- Fig. S2 Flow of the prognostic cohort.
- Fig. S3 Bland–Altman plots of sarco and TotalSegmentator on the reference L3 slice.
- Fig. S4 Muscle and marrow age.
- Table S2 Scanners by dataset.
- Table S3 Agreement of the deployed (in-sample) models.
- Table S4 Results by slice thickness and craniocaudal extent, including TotalSegmentator.
- Table S5 Effect of level selection; within-examination reproducibility in Dataset L.
- Table S6 Fixed-threshold classification of low muscle attenuation.
- Table S7 Discrimination of all prognostic models; bootstrap CIs of ΔC.
- Table S8 Hazard ratios of the main predictors.
- Tables S9 and S10 CLAIM 2024 and STROBE checklists.
- Table S11 Coefficients of the phase-0 conversion.
- Table S12 Truncated scans: cross-fitted and deployed localisation models, with and without the edge rule.
- Table S13 Agreement of TotalSegmentator with the reference labels.
- Table S14 Prognostic analyses with phase-0 areas.
- Table S15 Contrast effect by dataset, phase and level (as measured; one examination per patient).
- Table S16 Accuracy of the phase-0 conversion of areas.

