## Supplementary for "Deep-learning body composition analysis of routine CT robust to scan range, slice order and contrast phase"

Japanese Journal of Radiology

Shouhei Hanaoka, Wataru Gonoi, Shohei Inui, Nobuhiko Akamatsu, Yukihiro Nomura, Tomomi Takenaga, Soichiro Miki, Takeharu Yoshikawa, Kotaro Sugawara, Satoru Taguchi, Kenjiro Kishitani, Haruki Kume, Taketo Kawai, Tohru Nakagawa, Osamu Abe

Reference numbers in square brackets refer to the reference list of the main text; references numbered S1–S10 are listed at the end of this document. Contrast phases are numbered 0 (non-contrast), 1 (early arterial), 2 (late arterial), 3 (portal venous), 4 (venous) and 5 (equilibrium).

### Contents


- Methods S1. Model details, training and orientation evaluation (Table S1)
- Methods S2. Height and weight estimation
- Methods S3. Secondary and sensitivity analyses
- Methods S4. Muscle and marrow age
- Methods S5. Comparison with TotalSegmentator
- Fig. S1. Series by slice thickness and craniocaudal extent, and whether L1/L3 were found
- Fig. S2. Flow of the prognostic cohort
- Fig. S3. Bland–Altman plots of sarco and TotalSegmentator on the reference L3 slice
- Fig. S4. Muscle and marrow age
- Table S2. Scanners by dataset
- Table S3. Agreement of the deployed (in-sample) models with the reference labels
- Table S4. Results by slice thickness and craniocaudal extent
- Table S5. Effect of level selection on the measures, and within-examination reproducibility
- Table S6. Classification of low muscle attenuation with fixed thresholds as measured and after conversion
- Table S7. Discrimination of all prognostic models and bootstrap CIs of ΔC
- Table S8. Hazard ratios of the main predictors
- Table S9. CLAIM 2024 checklist
- Table S10. STROBE checklist (prognostic analysis)
- Table S11. Coefficients of the phase-0 conversion
- Table S12. Truncated scans: cross-fitted and deployed localisation models, with and without the edge rule
- Table S13. Agreement of TotalSegmentator with the reference labels
- Table S14. Prognostic analyses with phase-0 areas
- Table S15. Contrast effect by dataset, phase and level (as measured; one examination per patient)
- Table S16. Accuracy of the phase-0 conversion of areas
- References S1–S10


---

### Methods S1. Model details and training

#### Table S1. Models in sarco v8.6.0

| Component | Architecture and input | Training data | Training | Output and decision rule |
|---|---|---|---|---|
| Orientation | Small CNN; coronal and sagittal mean and maximum intensity projections on a 5 mm × 4 mm grid, in stored slice order | Datasets L, main, HC and P (train 3076, validation 647, test 671 series) | Random craniocaudal flips; random crops of 100–640 mm placed at a random position of the input window; narrowed field of view in half of the samples | Reverse the volume if P(feet first) > 0.7 |
| L1/L3 localisation | U-Net on frontal and lateral maximum and mean intensity projections; likelihood of the level per craniocaudal position | Dataset main (training and validation splits); version 4 adds Dataset HC series whose deployed result had been visually confirmed (L1 297, L3 318 series) | Fine-tuned from an earlier model trained on all of Dataset main: 300 epochs with random craniocaudal crops (probability 0.6; 120 mm to the whole volume), then 300 epochs with Dataset HC added; Dice and binary cross-entropy loss | Level absent if peak likelihood < threshold chosen on validation data (L1 0.303, L3 0.258) or within 10 mm of a scan end |
| Segmentation | Two-dimensional U-Net (4 output classes; 16 base channels); one slice at 1 mm pixels, 512 × 512, trunk extracted | Dataset main (all 2039 series; cross-fitting: four of five folds) | 1000 epochs; Dice and cross-entropy loss; Adam (learning rate $10^{-3}$, $\beta_1$ 0.99); batch size 40; no validation set; no augmentation | Muscle, cortical bone, medullary bone |
| Contrast phase | 2.5-dimensional ResNet18 with CBAM (ct25d-regressor); three 5-mm slabs at 5-mm intervals around the level; visceral region mask, 512 px × 1.0 mm; signed-distance mask channel; CT window −1000 to 1000 HU | Datasets L, P, HC (≤ 100 patients) and O; patient-level 70/15/15 split (seed 0) | Cross-entropy; temperature scaling on validation data | Six classes; probabilities of available levels averaged |
| Phase-0 conversion | Attenuation and proportions: linear, $v_0 = a + b \times v_k$; areas (SMA, VAT, SAT; from v8.6): power, $v_0 = c \times v_k^{\,p}$ ($v_k$, value in phase $k$; $v_0$, non-contrast equivalent); per level, measure and phase | Same-examination pairs (phase 1, Datasets L and O; 2, L and O; 3, L; 4, O and P; 5, L and P); released equations fitted on all splits, evaluation equations on the training and validation splits only; coefficients in Table S11 | Least squares (linear); weighted non-linear least squares with variance proportional to area (power) | Equation of the estimated phase; proportions clipped to 0–1 |
| Age (4 models) | As for contrast phase; vertebral medulla (256 px × 0.78125 mm) or muscle (512 px × 1.0 mm), L1 or L3; distance gate of 1 cm around the structure | Datasets L and main; patient-level 70/15/15 split | Gaussian output (mean and variance): Huber loss, then heteroscedastic and β-negative log-likelihood; ±20 HU intensity shift per case | Inverse-variance combination; width of the 95% interval scaled on validation data (factor 1.67–1.80) |

CNN, convolutional neural network; CBAM, convolutional block attention module. The age models used in the prognostic analysis (version 10) were trained in the same way on Datasets L, P, HC and O only, without Dataset main. The phase and conversion models did not use Dataset main. Orientation: 16 series with inconsistent level labels (14 Dataset main, 2 Dataset L) were excluded (3076 + 647 + 671 + 16 = 4410).

**Cross-fitting.** Dataset main was divided into five folds (stratified by cancer type and cancer-specific death; 408/408/408/408/407 series). For test fold k, the localisation model was trained from random initialisation on three folds with fold (k + 1) mod 5 for validation (600 epochs, then 300 epochs with the Dataset HC series), and the segmentation U-Net on the remaining four folds (1619–1621 series: 1631–1632 series minus the 2–3 series per fold with inconsistent reference labels) with the deployed settings. On validation data, the localisation models of all folds detected the presence of the level with an accuracy of 0.995–1.000 and placed it within 10 mm in 0.958–0.976 (L1) and 0.963–0.978 (L3) of series. The cross-fitted pipeline processed all 2039 series without failure; no series was reversed by the orientation step.

**Software.** sarco v8.6.0 (commit d06ec42; Zenodo https://doi.org/10.5281/zenodo.24158133). The analyses were run with v8.5.0 (commit 283f552), which uses the same models; v8.6.0 adds the phase-0 equivalent areas, which were computed with v8.6.0, and gave identical outputs otherwise when run in the same computing environment (160 series of the five datasets, including 10 cross-fitted series, compared). Other software: ct25d-regressor (commit 0f3b7b1); Python 3.12; PyTorch 2.11; statsmodels 0.15.0. Processing time: median 62.7 s (interquartile range 59.7–64.6 s; range 54.5–79.6 s) per series for 20 randomly selected series of Dataset main processed one at a time on one NVIDIA RTX PRO 6000 Blackwell Max-Q GPU with 32 CPU cores, from reading the volume to writing all results, without cached preprocessing. TotalSegmentator 2.18.0 with nnU-Net v2 2.8.1; weights: `total` 1.5 mm (nnU-Net task identifiers 291–295; vertebrae in 292), `total` 3 mm (task 297; phase estimation) and `tissue_types` (task 481).

**Orientation evaluation.** The orientation classifier was evaluated on the test splits, uncut and cropped to 200 mm (Dataset L) or 300 mm (Dataset main), and compared with an earlier version trained only on abdominal CT of Datasets L and main, with random craniocaudal crops of 100–640 mm but without narrowing of the field of view or random placement of the crop in the input window. The training of the current classifier used random flips, random craniocaudal crops of 100–640 mm and a narrowed field of view in half of the samples. Results are given in the main text (Results, Orientation).

### Methods S2. Height and weight estimation

Height and weight were estimated from projections of the trunk (arms and table removed; maximum and mean intensity projections in two directions at 2 mm, in coordinates relative to L3) and the L1/L3 axial slices, by two ResNet-CBAM branches with Gaussian heads. Augmentation included craniocaudal cropping that kept L1–L3, a narrowed reconstruction field of view (radius 175–250 mm) and dropping one level (probability 0.15). Sex and age were not used as inputs. On the held-out test split of Dataset main, the MAE was 3.48 cm for height and 2.65 kg for weight (coverage of the 95% interval 0.94 and 0.92); with L1 only, 4.16 cm and 3.40 kg; with L3 only, 4.24 cm and 3.17 kg. A model trained without Dataset main had an MAE of 6.22 cm ($r = 0.42$) and 3.87 kg ($r = 0.90$) on all of Dataset main. Measured height was used for SMI in all analyses.

### Fig. S1. Series by slice thickness and craniocaudal extent

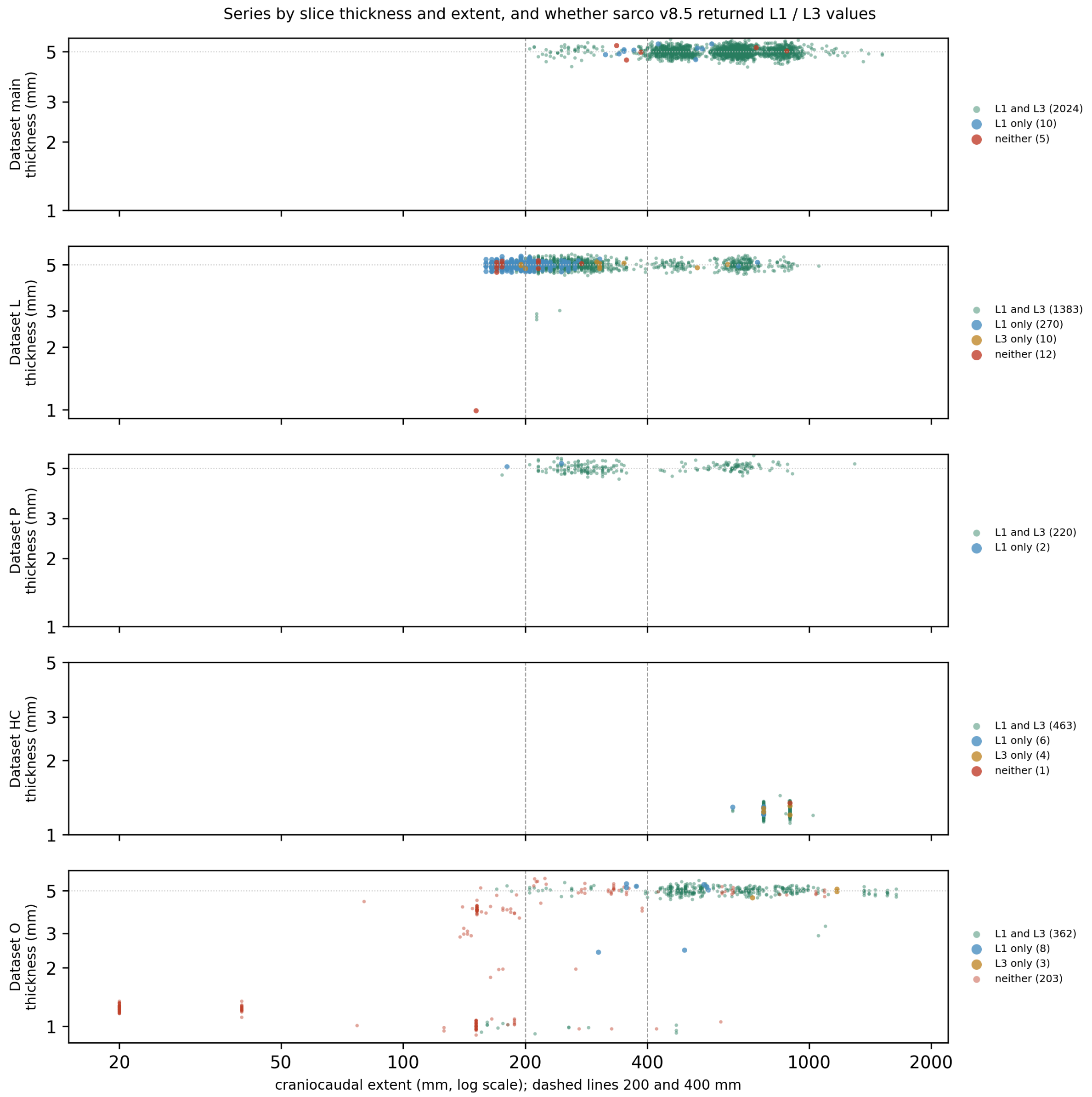


Craniocaudal extent (log scale) and slice thickness of every series of the five datasets, coloured by whether sarco returned L1 and L3, L1 only, L3 only or neither. Dashed lines, 200 and 400 mm.

Counts (series): both / L1 only / L3 only / neither

| Dataset | Extent < 200 mm | 200–400 mm | ≥ 400 mm |
|---|---|---|---|
| main | – | 79 / 5 / 0 / 3 (n = 87) | 1945 / 5 / 0 / 2 (n = 1952) |
| L | 105 / 143 / 2 / 8 (n = 258) | 1000 / 125 / 6 / 4 (n = 1135) | 278 / 2 / 2 / 0 (n = 282) |
| P | 1 / 1 / 0 / 0 (n = 2) | 123 / 1 / 0 / 0 (n = 124) | 96 / 0 / 0 / 0 (n = 96) |
| HC | – | – | 463 / 6 / 4 / 1 (n = 474) |
| O | 11 / 0 / 0 / 149 (n = 160) | 27 / 4 / 0 / 34 (n = 65) | 324 / 4 / 3 / 20 (n = 351) |

### Fig. S2. Flow of the prognostic cohort

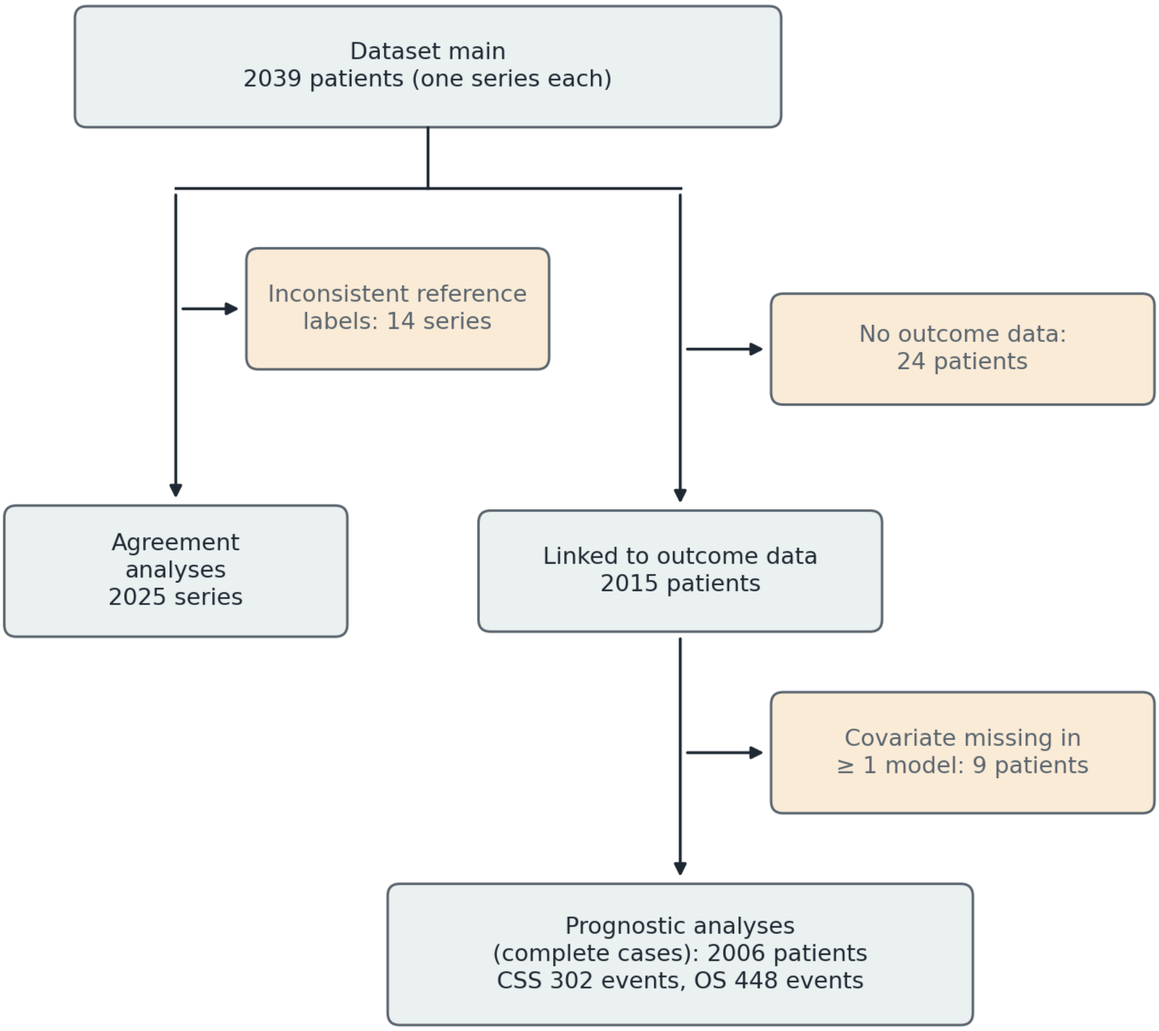


Flow of Dataset main into the agreement and prognostic analyses.

Dataset main, 2039 series ⟶ linked to the outcome data of the published cohort [4]: 2015 patients (24 series without outcome data) ⟶ complete data for the fully automated analysis: 2006 patients (9 excluded because a covariate of at least one model was missing, e.g. a level not found; one complete-case set was used for all models) ⟶ CSS 302 events, OS 448 events. Portal-venous-only subgroup (estimated phase): 1163 patients. Sensitivity analysis excluding the four patients who also appeared in the training data of the age or phase models: 2002 patients (302 CSS events).

Patients in more than one dataset: Dataset main shared 2, 1, 1 and 3 patients with Datasets L, P, HC and O, respectively.

For the agreement analyses, 14 of the 2039 series with inconsistent reference labels (e.g. swapped L1/L3 labels or joined volumes) were excluded (2025 series); they were not excluded from the prognostic analysis, which used only the pipeline's own measurements.

### Table S2. Scanners by dataset

| | Dataset main | Dataset L | Dataset P | Dataset HC | Dataset O |
|---|---|---|---|---|---|
| Unit (n) | Series (2039) | Series (1675) | Examinations (95; 94 patients) | Series (446) | Examinations (139; 134 patients) |
| Years | 2005–2018 | 2022 | 2025 | 2007–2008 | 2025 |
| Canon (Toshiba) | 1550 | 1167 | 55 | 0 | 89 |
| GE | 444 | 508 | 40 | 446 | 49 |
| Siemens | 16 | 0 | 0 | 0 | 1 |
| Philips | 15 | 0 | 0 | 0 | 0 |
| Hitachi | 1 | 0 | 0 | 0 | 0 |
| Unknown | 13 | 0 | 0 | 0 | 0 |

Models: Dataset main — Canon Aquilion 687, Aquilion ONE 553, Aquilion PRIME 207, Aquilion Precision 83, Aquilion/LB 17, Aquilion Prime SP 2, Aquiduo 1; GE Discovery CT750 HD 317, LightSpeed VCT 113, LightSpeed Ultra 5, Optima CT660 5, BrightSpeed 3, LightSpeed QX/i 1; Siemens Emotion 16 9, SOMATOM Definition AS+ 3, SOMATOM Definition 2, SOMATOM Definition AS 1, Sensation 64 1; Philips Brilliance 16 13, Brilliance 64 1, IQon Spectral CT 1; Hitachi SCENARIA 1; unknown 13 (workstation reconstructions). Dataset L — Aquilion ONE 771, Aquilion PRIME 396, Discovery CT750 HD 508. Dataset P — Aquilion ONE 21, Aquilion PRIME 27, Aquilion Prime SP 7, Discovery CT750 HD 16, Revolution CT 24. Dataset HC — Discovery STE (CT of PET/CT). Dataset O — Aquilion ONE 23, Aquilion PRIME 31, Aquilion Prime SP 35, Discovery CT750 HD 21, Revolution CT 27, Discovery 690 1, Biograph64 Vision 600 1. Vendor records were available for 94 of 99 (P), 446 of 474 (HC) and 134 of 144 (O) patients; in Datasets P and O, records were per examination.

### Table S3. Agreement of the deployed models (in sample) with the reference labels

The deployed segmentation and localisation models were trained on all of Dataset main; these values are therefore in-sample and given for reference only.

| | L1 | L3 |
|---|---|---|
| (a) Dice muscle / cortical / medullary | 0.973 / 0.891 / 0.973 | 0.974 / 0.885 / 0.976 |
| (a) SMA ICC(A,1) [95% CI]; ICC(C,1) [95% CI] | 0.996 [0.626–0.999]; 0.999 [0.999–0.999] | 0.999 [0.985–1.000]; 1.000 [0.999–1.000] |
| (a) SMA bias [LoA], $cm^2$ | +1.81 [0.25, 3.36] | +0.91 [−0.62, 2.43] |
| (a) MSMD ICC(A,1); bias [LoA], HU | 0.999; −0.38 [−0.85, 0.09] | 1.000; −0.18 [−0.56, 0.21] |
| (b) Slice same / ±1 / further / not found | 1991 / 5 / 22 / 2 | 2007 / 1 / 2 / 0 |
| (b) SMA ICC(A,1) [95% CI]; ICC(C,1) | 0.995 [0.806–0.999]; 0.999 | 0.999 [0.988–1.000]; 1.000 |
| (b) SMA bias [LoA], $cm^2$ | +1.80 [−0.31, 3.92] | +0.90 [−0.74, 2.53] |
| (b) SMI ICC(A,1) [95% CI]; ICC(C,1) | 0.993 [0.742–0.998]; 0.998 | 0.998 [0.983–0.999]; 0.999 |
| (b) MSMD ICC(A,1); ICC(C,1) | 0.998; 0.999 | 1.000; 1.000 |
| (b) $\|\Delta SMA\| > 10$ $cm^2$ | 4 | 1 |
| Low muscle mass (Fujiwara): agreement, κ | – | 0.988, 0.956 (n = 2001) |

The wide CIs of ICC(A,1) arise from a small constant bias combined with a very small random error; ICC(C,1) is not affected [25].

### Table S4. Results by slice thickness and craniocaudal extent

Strata were fixed before the analysis.

#### (a) Contrast-phase estimation (409 series assessed by both methods)

| Stratum | n | Four groups, sarco | Four groups, TotalSegmentator | Six phases, sarco | Equilibrium (n) |
|---|---|---|---|---|---|
| Thickness < 5 mm | 73 | 0.986 | 1.000 | 0.973 | 0 |
| Thickness ≥ 5 mm | 336 | 0.940 | 0.750 | 0.866 | 55 |
| Extent < 200 mm | 42 | 0.952 | 0.643 | 0.905 | 12 |
| Extent 200–400 mm | 199 | 0.960 | 0.668 | 0.915 | 42 |
| Extent ≥ 400 mm | 168 | 0.935 | 0.982 | 0.845 | 1 |
| Dataset L, thickness ≥ 5 mm | 257 | 0.953 | 0.700 | 0.930 | 51 |
| Dataset HC, thickness < 5 mm | 70 | 1.000 | 1.000 | 1.000 | 0 |
| Dataset P, thickness ≥ 5 mm | 34 | 0.971 | 0.882 | 0.794 | 4 |
| Dataset O, thickness ≥ 5 mm | 45 | 0.844 | 0.933 | 0.556 | 0 |
| Dataset O, thickness < 5 mm | 3 | 0.667 | 1.000 | 0.333 | 0 |

#### (b) L3 against the reference slice, each tool with its own slice (Dataset main; all series ≥ 5 mm)

| Stratum | Tool | n | Found | Same slice | ≤ 5 mm | ≤ 10 mm | SMA difference, mean (SD), cm² | \|ΔSMA\| > 10 cm² |
|---|---|---|---|---|---|---|---|---|
| Extent 200–400 mm | TotalSegmentator | 79 | 79 | 13 | 38 | 56 | −8.81 (7.11) | 37 |
| Extent 200–400 mm | sarco | 79 | 79 | 58 | 76 | 76 | +1.41 (3.48) | 2 |
| Extent ≥ 400 mm | TotalSegmentator | 1931 | 1911 | 221 | 898 | 1492 | −8.11 (9.87) | 819 |
| Extent ≥ 400 mm | sarco | 1931 | 1928 | 1405 | 1836 | 1844 | +1.35 (4.52) | 87 |

No series of Dataset main was thinner than 5 mm or shorter than 200 mm.

### Table S5. Effect of level selection on the measures, and within-examination reproducibility

#### (a) Dataset main, cross-fitted pipeline: difference from the reference (pipeline − reference) by slice-shift group

| Level | Group | n (%) | SMA, mean / SD [LoA], cm² | MSMD, mean / SD [LoA], HU | Usable* |
|---|---|---|---|---|---|
| L1 | Same slice | 1474 (73.0) | +0.80 / 3.83 [−6.7, 8.3] | −0.20 / 0.81 [−1.8, 1.4] | 1434 |

| Level | Group | n (%) | SMA, mean / SD [LoA], cm² | MSMD, mean / SD [LoA], HU | Usable* |
|---|---|---|---|---|---|
| L1 | ±1 slice | 459 (22.7) | +0.68 / 4.43 [−8.0, 9.4] | −0.02 / 1.68 [−3.3, 3.3] | 438 |
| L1 | ≥ 2 slices, same vertebra | 23 (1.1) | +1.47 / 5.65 [−9.6, 12.6] | +0.60 / 2.49 [−4.3, 5.5] | 20 |
| L1 | Other vertebra | 59 (2.9) | +0.82 / 12.09 [−22.9, 24.5] | −1.09 / 4.22 [−9.4, 7.2] | 28 |
| L1 | Not found | 5 (0.2) | – | – | – |
| L3 | Same slice | 1463 (72.8) | +1.26 / 3.64 [−5.9, 8.4] | −0.18 / 0.66 [−1.5, 1.1] | 1430 |
| L3 | ±1 slice | 449 (22.3) | +1.72 / 5.17 [−8.4, 11.9] | −0.13 / 1.47 [−3.0, 2.8] | 419 |
| L3 | ≥ 2 slices, same vertebra | 9 (0.4) | +0.58 / 8.92 [−16.9, 18.1] | +0.46 / 1.82 [−3.1, 4.0] | 6 |
| L3 | Other vertebra | 86 (4.3) | +1.04 / 9.88 [−18.3, 20.4] | +0.67 / 3.55 [−6.3, 7.6] | 50 |
| L3 | Not found | 3 (0.1) | – | – | – |

*Usable: level found with |ΔSMA| ≤ 10 cm² and |ΔMSMD| ≤ 5 HU (defined before the analysis). Overall usable: L1 1920/2020 (95.0%), L3 1905/2010 (94.8%). "Other vertebra": shift ≥ half the median vertebral height (16 mm; the median vertebral height, taken as half the L1–L3 distance of the reference labels in Dataset main, was 32.5 mm).

**(b) Dataset L: positions chosen in two series of the same examination with different phases**

| | Pairs compared | Identical slice geometry | Of these ≤ 10 mm | All pairs ≤ 10 mm | > 100 mm (coordinate mismatch) |
|---|---|---|---|---|---|
| L1, all series | 2918 | 1102 | 1066 (96.7%) | 2050 (70.3%) | 612 |
| L3, all series | 2228 | 822 | 794 (96.6%) | 1536 (68.9%) | 496 |
| L1, test split | 455 | 164 | 160 (97.6%) | 289 (63.5%) | 117 |
| L3, test split | 368 | 134 | 134 (100%) | 245 (66.6%) | 95 |

Pairs with different slice geometry often had inconsistent coordinate origins after conversion, so only pairs with identical geometry are interpretable.

### Table S6. Classification of low muscle attenuation with fixed thresholds

Thresholds of Sugawara et al. [5] (from Derstine et al. [13]): L3 MSMD < 38.5 HU (men) and < 34.3 HU (women). Dataset main, cross-fitted pipeline, 2010 patients: the patients of the prognostic cohort (2015 with outcome data) for whom the cross-fitted pipeline gave an L3 MSMD and sex was known. The hazard ratios below were estimated in the same 2010 patients, none of whom lacked any of the four covariates. The prognostic analysis in the main text used the 2006 patients with complete data for the covariates of all models.

| Estimated phase | n | As measured, low | Phase-0 equivalent, low | Portal-venous equivalent, low |
|---|---|---|---|---|
| All | 2010 | 442 (22.0%) | 1137 (56.6%) | 436 (21.7%) |
| Non-contrast | 11 | 8 | 8 | 4 |
| Early arterial | 54 | 31 (57.4%) | 37 | 16 (29.6%) |
| Late arterial | 140 | 41 (29.3%) | 79 | 30 (21.4%) |
| Portal venous | 1165 | 226 (19.4%) | 610 (52.4%) | 226 (19.4%) |
| Venous | 238 | 53 | 131 | 56 |
| Equilibrium | 402 | 83 (20.6%) | 272 | 104 (25.9%) |

| | n low | HR for OS [95% CI] | HR for CSS [95% CI] |
|---|---|---|---|
| As measured | 442 | 1.50 [1.21–1.85] | 1.27 [0.96–1.68] |
| Phase-0 equivalent | 1137 | 1.18 [0.97–1.44] | 1.08 [0.85–1.37] |
| Portal-venous equivalent | 436 | 1.40 [1.13–1.74] | 1.23 [0.93–1.62] |

Adjusted for age ≥ 70 years, sex, urological cancer and stage IV. Published prevalence in the same cohort: 423/2015 (21.0%) [5]. Applying the original thresholds to portal-venous-equivalent values corresponds to phase-0 thresholds of 31.2 HU (men) and 27.4 HU (women). In Dataset L (test split, 162 contrast series paired with non-contrast series of the same examination), disagreement of the classification with the non-contrast series decreased from 12/162 (7.4%) as measured to 7/162 (4.3%) on the portal-venous-equivalent scale, and false "low" in the early arterial phase from 5/23 (22%) to 0/23.

### Table S7. Discrimination of all prognostic models (fully automated, n = 2006)

Uno's C at 60 months, apparent / cross-validated (10-fold × 20 repeats). Values with the reference labels in parentheses (apparent / cross-validated).

| Model | Covariates | CSS | OS |
|---|---|---|---|
| A0 | Age ≥ 70, sex, urological cancer, stage IV | 0.661 / 0.648 (0.662 / 0.652) | 0.671 / 0.665 (0.672 / 0.667) |
| A1 | A0 + low muscle mass (Fujiwara) | 0.693 / 0.682 (0.692 / 0.683) | 0.701 / 0.696 (0.708 / 0.701) |
| A2 | A1 + phase-0 L3 MSMD | 0.695 / 0.683 (0.697 / 0.684) | 0.706 / 0.700 (0.713 / 0.706) |
| A5 | A2 + muscle and marrow age gaps | 0.696 / 0.679 (0.706 / 0.685) | 0.706 / 0.697 (0.715 / 0.707) |
| C0 | A0 + BMI | 0.696 / 0.682 (0.698 / 0.682) | 0.708 / 0.700 (0.709 / 0.701) |
| C1 | C0 + low muscle mass | 0.703 / 0.685 (0.703 / 0.684) | 0.715 / 0.707 (0.718 / 0.709) |
| C2 | C1 + phase-0 L3 MSMD | 0.711 / 0.683 (0.712 / 0.685) | 0.727 / 0.713 (0.729 / 0.714) |
| C3 | C2 + age gaps | 0.711 / 0.680 (0.714 / 0.684) | 0.726 / 0.711 (0.730 / 0.714) |
| C4 | C0 + phase-0 L3 MSMD | 0.709 / 0.687 (0.710 / 0.686) | 0.724 / 0.712 (0.725 / 0.713) |
| C11 | C2 + phase-0 L3 subcutaneous fat attenuation | 0.719 / 0.693 (0.715 / 0.691) | 0.734 / 0.718 (0.735 / 0.719) |

**Bootstrap 95% CIs of the apparent ΔC** (1000 patient-level resamples, models refitted, percentile method; fully automated | reference labels)

| Comparison | CSS | OS |
|---|---|---|
| A1 − A0 (primary) | +0.032 [0.010, 0.050] \| +0.029 [0.009, 0.048] | +0.030 [0.017, 0.045] \| +0.036 [0.020, 0.051] |
| C0 − A0 | +0.036 [0.011, 0.063] | +0.037 [0.020, 0.058] |
| C4 − C0 (primary for OS) | +0.012 [0.001, 0.028] \| +0.013 [0.000, 0.029] | +0.017 [0.007, 0.029] \| +0.015 [0.006, 0.029] |
| C2 − C1 | +0.008 [−0.002, 0.024] | +0.012 [0.004, 0.023] |
| C3 − C2 (age gaps) | 0.000 [−0.004, 0.009] | −0.001 [−0.002, 0.004] |
| C11 − C2 | +0.007 [−0.002, 0.025] | +0.007 [0.002, 0.017] |

Reference-label values are given for the primary comparisons only. Likelihood-ratio tests of the nested models (fully automated) gave P < 0.001 for all comparisons above except C3 − C2 (CSS P = 0.998; OS P = 0.80). Apparent ΔC of nested models is optimistic; the cross-validated ΔC is the primary measure (main text). Results were similar when the four patients who were also in the training data of the age or phase models were excluded (apparent C for CSS: A1 0.692, C4 0.709). Exploratory: all comparisons other than A1 − A0 and C4 − C0.

### Table S8. Hazard ratios of the main predictors (fully automated; reference labels in parentheses)

| Model: predictor | CSS, HR [95% CI] | OS, HR [95% CI] |
|---|---|---|
| A1: low muscle mass (Fujiwara) | 1.88 [1.43–2.47] (1.77 [1.36–2.30]) | 2.32 [1.88–2.86] (2.23 [1.82–2.73]) |
| A2: phase-0 L3 MSMD, per 10 HU | 0.87 [0.74–1.03] (0.90 [0.77–1.05]) | 0.81 [0.71–0.93] (0.82 [0.72–0.93]) |
| C4: BMI, per 5 kg/m² | 0.54 [0.44–0.67] (0.55) | 0.44 [0.37–0.52] (0.44) |
| C4: phase-0 L3 MSMD, per 10 HU | 0.70 [0.59–0.83] (0.73 [0.61–0.86]) | 0.61 [0.53–0.70] (0.62 [0.54–0.71]) |
| C2: low muscle mass | 1.28 [0.93–1.76] (1.24 [0.91–1.69]) | 1.41 [1.10–1.81] (1.39 [1.09–1.76]) |
| C2: phase-0 L3 MSMD, per 10 HU | 0.72 [0.60–0.87] (0.75) | 0.64 [0.55–0.74] (0.65) |
| C3: muscle age gap, per 5 years | 1.00 [0.90–1.10] (0.95) | 1.03 [0.95–1.12] (0.98) |
| C3: marrow age gap, per 5 years | 1.00 [0.93–1.07] (1.02) | 1.00 [0.94–1.06] (1.03) |
| C11: phase-0 L3 subcutaneous fat attenuation, per 10 HU | 1.36 [1.21–1.54] (1.35 [1.20–1.52]) | 1.28 [1.16–1.42] (1.27 [1.15–1.41]) |

Portal-venous-only subgroup (estimated phase, n = 1163): A1 low muscle mass HR 1.81 (CSS) and 2.09 (OS); C4 MSMD HR 0.77 (CSS) and 0.71 (OS).

---

### Table S9. CLAIM 2024 checklist

| # | Item | Location |
|---|---|---|
| 1 | Identification as an AI study | Title ("deep-learning"); Abstract (U-Net); Keywords |
| 2 | Structured abstract | Abstract |
| 3 | Background, intended use and clinical role | Introduction |
| 4 | Objectives and hypotheses | Introduction, last paragraph |
| 5 | Prospective or retrospective | Methods, Ethics and study design |
| 6 | Study goal | Methods, Ethics and study design; Introduction |
| 7 | Data sources | Methods, Datasets; Table 1 |
| 8 | Eligibility criteria | Methods, Datasets; Dataset main as in [4] |
| 9 | Data pre-processing | Methods, Pipeline; Methods S1 |
| 10 | Selection of data subsets | Methods, Pipeline (L1/L3 slices); Evaluation |
| 11 | De-identification | Methods, Datasets |
| 12 | Missing data | Methods, Prognosis; Fig. S2 |
| 13 | Image acquisition | Table 1; Table S2 |
| 14 | Reference standard definition | Methods, Reference labels; Datasets (reference phase) |
| 15 | Rationale and limitations of the reference standard | Methods, Reference labels; Discussion |
| 16 | Annotators and qualifications | Methods, Reference labels; as described in [4] |
| 17 | Annotation methods and tools | Methods, Reference labels (ITK-SNAP, CIRCUS [15]) |
| 18 | Inter- and intra-rater variability | Not assessed (Discussion, limitations) |
| 19 | Data partitions | Methods, Evaluation (70/15/15; five-fold cross-fitting); Methods S1 |
| 20 | Level of partitioning | Patient level (Methods, Evaluation) |

| # | Item | Location |
|---|---|---|
| 21 | Test set size | Determined by the available data; no power calculation (Methods, Datasets) |
| 22 | Model architecture | Methods, Pipeline; Fig. 1; Table S1 |
| 23 | Software and versions | Methods, Statistics; Methods S1 |
| 24 | Initialisation of parameters | Methods S1 (cross-fitting from random initialisation; deployed localisation model fine-tuned) |
| 25 | Training details and hyperparameters | Table S1 |
| 26 | Model selection | Table S1 (validation thresholds; fixed epochs) |
| 27 | Ensembling | Methods S4 (inverse-variance combination of four age models) |
| 28 | Performance metrics | Methods, Evaluation |
| 29 | Statistical significance and uncertainty | Methods, Prognosis and Statistics; Tables 2 and 4; Table S7; no CIs for phase accuracy |
| 30 | Robustness and sensitivity analyses | Truncated scans (Fig. 3; Table S12); strata (Table S4); Methods S3 |
| 31 | Explainability | Not applicable; outputs are measurable quantities shown as overlays (Fig. 2) |
| 32 | Internal test performance and training performance | Results; Table S3 |
| 33 | External testing | Not performed (Discussion, limitations) |
| 34 | Trial registration | Not applicable; not pre-registered (Methods) |
| 35 | Flow of participants | Fig. S2 |
| 36 | Demographic and clinical characteristics | Table 1; [4] |
| 37 | Performance across subgroups | Tables 2 and 3; Table S4 |
| 38 | Estimates of diagnostic accuracy and precision | Table 3; Fig. 5 (accuracy without CIs) |
| 39 | Failure analysis | Fig. 5; Fig. 4; Results (failure types); Table S5 |
| 40 | Limitations | Discussion |
| 41 | Implications for practice | Discussion |
| 42 | Access to full protocol | No separate protocol; the analysis specification is given in Methods (Ethics and study design; Evaluation) and Methods S1–S3 |
| 43 | Availability of software, models and data | Data availability |
| 44 | Funding | Declarations (funders had no role) |

### Table S10. STROBE checklist (prognostic analysis; cohort study)

| # | Item | Location |
|---|---|---|
| 1 | Title and abstract | Title; Abstract |
| 2 | Background and rationale | Introduction |
| 3 | Objectives | Introduction (aim 4) |
| 4 | Study design | Methods, Ethics and study design |
| 5 | Setting, dates, follow-up | Methods, Datasets and Prognosis (surgery 2006–2018 [4]; follow-up to December 2022) |
| 6 | Participants | Methods, Datasets; [4] |
| 7 | Variables | Methods, Prognosis |

| # | Item | Location |
|---|---|---|
| 8 | Data sources and measurement | Methods, Pipeline and Prognosis |
| 9 | Bias | Methods, Cross-fitting; Discussion |
| 10 | Study size | All patients of the existing cohort (Fig. S2) |
| 11 | Quantitative variables | Methods, Prognosis (per 10 HU; Fujiwara criteria [23]) |
| 12 | Statistical methods | Methods, Prognosis and Statistics; Table S7 |
| 13 | Participants at each stage | Fig. S2 |
| 14 | Descriptive data | Table 1; [4] |
| 15 | Outcome data | Results; Table 4 (302 and 448 events) |
| 16 | Main results | Table 4; Table S8 |
| 17 | Other analyses | Tables S6–S8 |
| 18 | Key results | Discussion, first paragraph |
| 19 | Limitations | Discussion |
| 20 | Interpretation | Discussion |
| 21 | Generalisability | Discussion (single centre) |
| 22 | Funding | Declarations (funders had no role) |

### Methods S3. Secondary and sensitivity analyses

All analyses below are exploratory and used the same 2006 patients, folds and Cox models as the primary analyses unless stated otherwise.

- **Muscle and marrow age gaps:** residuals of the predicted age regressed on chronological age [S6, S7], computed with age models trained without Dataset main (Methods S4).
- **Additional models** (Table S7): A2 and C2 add phase-0 L3 MSMD to A1 and C1; A5 and C3 add the muscle and marrow age gaps; C11 adds phase-0 L3 subcutaneous fat attenuation to C2. Models with fat areas (Table S14): A6 and A7 add L3 VAT, L3 SAT and BMI to A5 and A1; C5 and C6 add the L3 VAT and SAT indices (area divided by height squared) to C2 and C3.
- **Nested-model tests:** likelihood-ratio tests of each added term.
- **Portal-venous-only subgroup:** patients whose series was estimated as portal venous (n = 1163); models A1 and C4 refitted.
- **Proportional hazards:** Schoenfeld residuals correlated with Kaplan–Meier-transformed time and with time rank, for each covariate of A0, A1, C0 and C4 (CSS and OS); P = 0.143–0.867 (CSS) and 0.147–0.718 (OS).
- **Phase-0 areas (Table S14):** SMA, VAT and SAT converted with the power equations; all models refitted.
- **Overlap sensitivity analysis:** excluding the four patients who were also in the training data of the age or phase models (n = 2002).
- **Fixed thresholds** (Table S6): portal-venous-equivalent values are phase-0 values converted back to the portal-venous scale with the inverse of the L3 MSMD equation for phase 3 ($v_0 = -3.62 + 0.904 \times v_3$); series estimated as portal venous keep their measured value. The original thresholds (38.5 and 34.3 HU) correspond to 31.2 and 27.4 HU on the phase-0 scale ($-3.62 + 0.904 \times 38.5$; $-3.62 + 0.904 \times 34.3$).

### Table S11. Coefficients of the phase-0 conversion

$v_0 = a + b \times v_k$. Pairs: same-examination pairs of a non-contrast series and a phase-$k$ series in all splits (released equations; the evaluation in Table 3 of the main text used equations refitted on the training and validation splits only). One examination may contribute more than one pair. The coefficients for all six measures (MSMD, VAT and SAT attenuation, cortical and medullary bone attenuation, and the proportion of muscle below −30 HU) are distributed with the model weights (`model/phase_correction.json`).

| Phase *k* | L3 *a* | L3 *b* | L3 pairs | L1 *a* | L1 *b* | L1 pairs |
|---|---|---|---|---|---|---|
| 1 Early arterial | −0.48 | 0.948 | 308 | −0.28 | 0.924 | 356 |
| 2 Late arterial | −1.80 | 0.905 | 322 | −2.20 | 0.883 | 369 |
| 3 Portal venous | −3.62 | 0.904 | 326 | −2.86 | 0.866 | 327 |
| 4 Venous | −4.34 | 0.914 | 176 | −2.13 | 0.851 | 174 |
| 5 Equilibrium | −4.01 | 0.869 | 292 | −3.90 | 0.868 | 343 |

Areas ($v_0 = c \times v_k{}^p$; areas in $cm^2$; weighted non-linear least squares with variance proportional to area; same pairs as above):

| Area | Phase *k* | L3 *c* | L3 *p* | L3 pairs | L1 *c* | L1 *p* | L1 pairs |
|---|---|---|---|---|---|---|---|
| SMA | 1 Early arterial | 1.334 | 0.939 | 308 | 0.931 | 1.012 | 356 |
| SMA | 2 Late arterial | 1.370 | 0.932 | 322 | 0.945 | 1.008 | 369 |
| SMA | 3 Portal venous | 1.216 | 0.956 | 326 | 0.920 | 1.013 | 327 |
| SMA | 4 Venous | 1.157 | 0.965 | 176 | 1.097 | 0.976 | 174 |
| SMA | 5 Equilibrium | 1.181 | 0.960 | 292 | 0.870 | 1.023 | 343 |
| VAT | 1 Early arterial | 1.619 | 0.919 | 308 | 1.471 | 0.936 | 356 |
| VAT | 2 Late arterial | 2.474 | 0.848 | 322 | 2.193 | 0.872 | 369 |
| VAT | 3 Portal venous | 2.630 | 0.838 | 326 | 2.472 | 0.852 | 327 |
| VAT | 4 Venous | 2.732 | 0.824 | 176 | 2.791 | 0.830 | 174 |
| VAT | 5 Equilibrium | 3.138 | 0.806 | 292 | 2.826 | 0.828 | 343 |
| SAT | 1 Early arterial | 1.096 | 0.983 | 308 | 1.059 | 0.989 | 356 |
| SAT | 2 Late arterial | 1.238 | 0.960 | 322 | 1.139 | 0.974 | 369 |
| SAT | 3 Portal venous | 1.375 | 0.939 | 326 | 1.214 | 0.963 | 327 |
| SAT | 4 Venous | 1.345 | 0.945 | 176 | 1.309 | 0.945 | 174 |
| SAT | 5 Equilibrium | 1.746 | 0.892 | 292 | 1.400 | 0.935 | 343 |

### Table S12. Truncated scans: cross-fitted and deployed localisation models, with and without the edge rule

In-range scenarios (a–c): proportion found / same slice as the reference / off by half a vertebra (16 mm) or more. Out-of-range scenarios (d, e): proportion with a false detection. Each volume was processed with the cross-fitted models that had not been trained on that patient. The deployed models were initialised from a model trained on all of Dataset main, so their results are in sample. The main text and Fig. 3 use the cross-fitted models with the edge rule.

| Level | Scenario | n | TotalSegmentator | Deployed, edge rule | Deployed, no rule | Cross-fitted, edge rule | Cross-fitted, no rule |
|---|---|---|---|---|---|---|---|
| L1 | (a) whole | 303 | 0.960 / 0.133 / 0.073 | 1.000 / 0.974 / 0.017 | 1.000 / 0.974 / 0.017 | 1.000 / 0.743 / 0.043 | 1.000 / 0.743 / 0.043 |
| L1 | (b) mid-range | 283 | 0.954 / 0.134 / 0.042 | 1.000 / 0.940 / 0.014 | 1.000 / 0.940 / 0.014 | 1.000 / 0.742 / 0.049 | 1.000 / 0.742 / 0.049 |
| L1 | (c) near end | 299 | 0.883 / 0.214 / 0.100 | 0.987 / 0.943 / 0.013 | 0.997 / 0.943 / 0.013 | 0.977 / 0.732 / 0.037 | 1.000 / 0.732 / 0.043 |
| L1 | (d) just outside | 297 | 0.104 | 0.084 | 0.229 | 0.071 | 0.330 |
| L1 | (e) far outside | 294 | 0.000 | 0.000 | 0.020 | 0.000 | 0.024 |
| L3 | (a) whole | 301 | 0.990 / 0.120 / 0.113 | 1.000 / 1.000 / 0.000 | 1.000 / 1.000 / 0.000 | 1.000 / 0.724 / 0.056 | 1.000 / 0.724 / 0.056 |
| L3 | (b) mid-range | 274 | 0.982 / 0.150 / 0.117 | 1.000 / 0.989 / 0.004 | 1.000 / 0.989 / 0.004 | 1.000 / 0.723 / 0.055 | 1.000 / 0.723 / 0.055 |
| L3 | (c) near end | 300 | 0.940 / 0.160 / 0.143 | 1.000 / 0.983 / 0.007 | 1.000 / 0.983 / 0.007 | 0.987 / 0.710 / 0.047 | 1.000 / 0.710 / 0.050 |
| L3 | (d) just outside | 295 | 0.136 | 0.078 | 0.210 | 0.132 | 0.261 |
| L3 | (e) far outside | 279 | 0.000 | 0.000 | 0.000 | 0.007 | 0.011 |

TotalSegmentator's L3 mask reached the scan end in 33 (L1) and 59 (L3) near-end volumes.

### Table S14. Prognostic analyses with phase-0 areas (sensitivity analysis)

SMA, VAT and SAT converted to phase-0 equivalents with the v8.6 power equations (Table S11); SMI, low muscle mass, VAT and SAT recomputed. All other settings as in the main analysis.

| Quantity | Areas as measured (primary) | Phase-0 areas |
|---|---|---|
| Low muscle mass, n of 2010 | 315 | 333 |
| Cross-validated C, A1, CSS / OS | 0.682 / 0.696 | 0.676 / 0.693 |
| Cross-validated ΔC A1 − A0, CSS [95% CI] | +0.034 [0.009, 0.053] | +0.028 [0.006, 0.047] |
| Cross-validated ΔC A1 − A0, OS [95% CI] | +0.031 [0.016, 0.048] | +0.028 [0.014, 0.043] |
| Apparent ΔC A1 − A0, CSS / OS [95% CI] | +0.032 [0.010, 0.050] / +0.030 [0.017, 0.045] | +0.027 [0.007, 0.044] / +0.028 [0.014, 0.042] |
| HR for low muscle mass (A1), CSS [95% CI] | 1.88 [1.43–2.47] | 1.73 [1.32–2.27] |
| HR for low muscle mass (A1), OS [95% CI] | 2.32 [1.88–2.86] | 2.10 [1.70–2.59] |
| C4 − C0 (L3 MSMD) | No area covariate | Unchanged |
| Models with VAT or SAT (A6, A7, C5, C6): change in cross-validated C | — | −0.001 to −0.004 |
| Proportional hazards (Schoenfeld residuals), smallest P | 0.143 | 0.143 |
| Reference labels: HR for low muscle mass, CSS / OS | 1.77 / 2.23 | 1.79 / 2.16 |

Fully automated (cross-fitted) cohort, 2006 patients with complete data (302 cancer-specific deaths; 448 deaths). Contrast-enhanced series, median change after conversion: L3 SMA −1.4%, VAT +28.6%, SAT

+4.7%. Cross-validated ΔC: mean of 20 repeats of 10-fold cross-validation (as in Table 4 of the main text), 95% CI from 500 patient-level bootstrap replicates of the whole cross-validation (5 repeats each); apparent ΔC: 1000 replicates. Reference labels: radiologist-corrected labels, 2002 patients.

### Table S15. Contrast effect by dataset, phase and level (as measured; one examination per patient)

#### (a) MSMD, HU

| Dataset | Phase | Level | Patients | Non-contrast, mean (SD) | Contrast-enhanced, mean (SD) | Difference, mean (SD) | All examinations: pairs | All examinations: difference, mean (SD) |
|---|---|---|---|---|---|---|---|---|
| L | Early arterial | L1 | 308 | 36.0 (7.9) | 39.1 (8.1) | +3.1 (2.2) | 321 | +3.1 (2.3) |
| L | Early arterial | L3 | 229 | 34.8 (8.6) | 36.9 (8.8) | +2.2 (1.9) | 239 | +2.2 (1.9) |
| L | Late arterial | L1 | 309 | 36.0 (7.9) | 43.5 (8.3) | +7.5 (2.5) | 322 | +7.5 (2.5) |
| L | Late arterial | L3 | 228 | 34.8 (8.6) | 40.7 (9.0) | +5.9 (2.7) | 236 | +5.9 (2.7) |
| L | Portal venous | L1 | 310 | 36.0 (7.9) | 44.9 (8.6) | +9.0 (2.9) | 324 | +9.0 (2.9) |
| L | Portal venous | L3 | 311 | 34.6 (8.5) | 42.2 (8.9) | +7.6 (2.6) | 325 | +7.6 (2.6) |
| L | Equilibrium | L1 | 306 | 35.9 (7.9) | 45.9 (8.6) | +9.9 (2.7) | 318 | +10.0 (2.7) |
| L | Equilibrium | L3 | 226 | 34.7 (8.7) | 44.5 (9.4) | +9.8 (2.9) | 234 | +9.8 (2.9) |
| P | Venous | L1 | 97 | 36.0 (9.2) | 44.6 (10.0) | +8.6 (3.3) | 98 | +8.6 (3.4) |
| P | Venous | L3 | 96 | 33.0 (9.7) | 41.1 (10.1) | +8.1 (3.0) | 97 | +8.1 (3.0) |
| P | Equilibrium | L1 | 23 | 35.9 (9.5) | 46.5 (9.6) | +10.6 (2.5) | 23 | +10.6 (2.5) |
| P | Equilibrium | L3 | 22 | 34.0 (10.8) | 44.3 (11.6) | +10.3 (2.9) | 22 | +10.3 (2.9) |

#### (b) Areas (SMA, VAT, SAT; cm²) and VAT and SAT attenuation (HU)

*SMA (cm²)*

| Dataset | Phase | Level | Patients | Non-contrast, mean (SD) | Contrast-enhanced, mean (SD) | Difference, mean (SD) | All examinations: pairs | All examinations: difference, mean (SD) |
|---|---|---|---|---|---|---|---|---|
| L | Early arterial | L1 | 308 | 101.8 (24.3) | 103.6 (24.0) | +1.8 (4.2) | 321 | +1.6 (4.2) |
| L | Early arterial | L3 | 229 | 107.0 (24.8) | 107.1 (25.8) | 0.0 (4.8) | 239 | +0.1 (4.8) |
| L | Late arterial | L1 | 309 | 101.8 (24.3) | 103.8 (24.2) | +2.0 (4.5) | 322 | +1.9 (4.5) |
| L | Late arterial | L3 | 228 | 106.5 (24.5) | 107.3 (25.6) | +0.8 (5.5) | 236 | +0.8 (5.5) |
| L | Portal venous | L1 | 310 | 102.3 (24.4) | 104.7 (24.3) | +2.5 (4.6) | 324 | +2.4 (4.6) |
| L | Portal venous | L3 | 311 | 110.3 (26.2) | 112.0 (27.3) | +1.7 (4.7) | 325 | +1.9 (4.8) |
| L | Equilibrium | L1 | 306 | 101.7 (24.2) | 104.9 (23.9) | +3.2 (4.7) | 318 | +3.2 (4.7) |
| L | Equilibrium | L3 | 226 | 106.4 (24.4) | 108.9 (25.3) | +2.5 (5.9) | 234 | +2.6 (6.0) |

| Dataset | Phase | Level | Patients | Non-contrast, mean (SD) | Contrast-enhanced, mean (SD) | Difference, mean (SD) | All examinations: pairs | All examinations: difference, mean (SD) |
|---|---|---|---|---|---|---|---|---|
| P | Venous | L1 | 97 | 97.9 (23.6) | 99.7 (24.4) | +1.8 (6.0) | 98 | +1.7 (6.0) |
| P | Venous | L3 | 96 | 102.1 (25.7) | 105.0 (26.6) | +2.9 (6.0) | 97 | +2.7 (6.2) |
| P | Equilibrium | L1 | 23 | 91.5 (22.9) | 93.7 (22.0) | +2.1 (4.3) | 23 | +2.1 (4.3) |
| P | Equilibrium | L3 | 22 | 98.7 (25.5) | 101.7 (25.3) | +3.0 (3.0) | 22 | +3.0 (3.0) |

#### *VAT area ($cm^2$)*

| Dataset | Phase | Level | Patients | Non-contrast, mean (SD) | Contrast-enhanced, mean (SD) | Difference, mean (SD) | All examinations: pairs | All examinations: difference, mean (SD) |
|---|---|---|---|---|---|---|---|---|
| L | Early arterial | L1 | 308 | 107.0 (83.0) | 99.9 (79.9) | −7.1 (11.1) | 321 | −7.2 (11.1) |
| L | Early arterial | L3 | 229 | 116.4 (87.0) | 109.1 (85.3) | −7.4 (8.9) | 239 | −7.5 (9.0) |
| L | Late arterial | L1 | 309 | 107.0 (82.9) | 90.0 (76.7) | −17.0 (13.1) | 322 | −17.0 (13.1) |
| L | Late arterial | L3 | 228 | 114.4 (84.9) | 98.3 (81.8) | −16.2 (11.0) | 236 | −16.2 (11.1) |
| L | Portal venous | L1 | 310 | 107.9 (83.5) | 89.4 (77.3) | −18.4 (13.3) | 324 | −18.5 (13.4) |
| L | Portal venous | L3 | 311 | 127.3 (87.6) | 108.7 (83.7) | −18.7 (11.2) | 325 | −18.9 (11.3) |
| L | Equilibrium | L1 | 306 | 106.9 (82.9) | 86.5 (76.0) | −20.4 (13.5) | 318 | −20.5 (13.6) |
| L | Equilibrium | L3 | 226 | 113.8 (83.3) | 92.9 (78.6) | −20.9 (12.3) | 234 | −20.9 (12.4) |
| P | Venous | L1 | 97 | 90.2 (66.0) | 67.2 (57.8) | −23.0 (20.7) | 98 | −22.8 (20.7) |
| P | Venous | L3 | 96 | 97.1 (68.2) | 81.6 (69.0) | −15.5 (14.0) | 97 | −15.5 (13.9) |
| P | Equilibrium | L1 | 23 | 86.6 (68.1) | 67.4 (58.9) | −19.2 (13.1) | 23 | −19.2 (13.1) |
| P | Equilibrium | L3 | 22 | 87.5 (54.9) | 72.4 (53.5) | −15.2 (6.2) | 22 | −15.2 (6.2) |

#### *SAT area ($cm^2$)*

| Dataset | Phase | Level | Patients | Non-contrast, mean (SD) | Contrast-enhanced, mean (SD) | Difference, mean (SD) | All examinations: pairs | All examinations: difference, mean (SD) |
|---|---|---|---|---|---|---|---|---|
| L | Early arterial | L1 | 308 | 77.9 (47.9) | 77.2 (47.7) | −0.7 (4.1) | 321 | −0.6 (4.2) |
| L | Early arterial | L3 | 229 | 106.7 (58.2) | 105.8 (57.5) | −0.9 (4.1) | 239 | −0.9 (4.1) |
| L | Late arterial | L1 | 309 | 77.8 (47.8) | 76.3 (47.6) | −1.5 (5.7) | 322 | −1.4 (5.7) |
| L | Late arterial | L3 | 228 | 105.7 (57.4) | 103.3 (57.4) | −2.5 (5.1) | 236 | −2.5 (5.1) |
| L | Portal venous | L1 | 310 | 78.0 (47.8) | 76.0 (47.7) | −2.0 (3.7) | 324 | −1.9 (3.8) |
| L | Portal venous | L3 | 311 | 112.0 (58.0) | 109.3 (58.6) | −2.7 (5.1) | 325 | −2.8 (5.2) |
| L | Equilibrium | L1 | 306 | 78.1 (47.8) | 74.4 (47.1) | −3.7 (4.5) | 318 | −3.7 (4.6) |

| Dataset | Phase | Level | Patients | Non-contrast, mean (SD) | Contrast-enhanced, mean (SD) | Difference, mean (SD) | All examinations: pairs | All examinations: difference, mean (SD) |
|---|---|---|---|---|---|---|---|---|
| L | Equilibrium | L3 | 226 | 106.6 (57.2) | 102.6 (57.7) | −4.0 (6.1) | 234 | −4.1 (6.1) |
| P | Venous | L1 | 97 | 68.9 (53.7) | 66.9 (53.5) | −1.9 (3.4) | 98 | −1.9 (3.4) |
| P | Venous | L3 | 96 | 102.0 (58.4) | 97.6 (55.9) | −4.4 (10.7) | 97 | −4.4 (10.6) |
| P | Equilibrium | L1 | 23 | 75.2 (58.7) | 72.0 (58.5) | −3.2 (2.0) | 23 | −3.2 (2.0) |
| P | Equilibrium | L3 | 22 | 110.7 (60.1) | 106.7 (60.5) | −4.0 (2.8) | 22 | −4.0 (2.8) |

#### *VAT attenuation (HU)*

| Dataset | Phase | Level | Patients | Non-contrast, mean (SD) | Contrast-enhanced, mean (SD) | Difference, mean (SD) | All examinations: pairs | All examinations: difference, mean (SD) |
|---|---|---|---|---|---|---|---|---|
| L | Early arterial | L1 | 308 | −87.7 (8.0) | −86.0 (7.9) | +1.7 (2.3) | 321 | +1.6 (2.6) |
| L | Early arterial | L3 | 229 | −88.2 (8.8) | −86.8 (8.6) | +1.4 (2.2) | 239 | +1.3 (2.3) |
| L | Late arterial | L1 | 309 | −87.7 (8.0) | −83.3 (7.7) | +4.4 (3.5) | 322 | +4.2 (4.0) |
| L | Late arterial | L3 | 228 | −88.1 (8.8) | −84.4 (8.2) | +3.7 (3.1) | 236 | +3.6 (3.2) |
| L | Portal venous | L1 | 310 | −87.7 (8.0) | −82.6 (7.6) | +5.1 (3.7) | 324 | +4.9 (4.2) |
| L | Portal venous | L3 | 311 | −88.9 (8.6) | −84.1 (7.7) | +4.8 (3.4) | 325 | +4.7 (3.5) |
| L | Equilibrium | L1 | 306 | −87.6 (8.0) | −82.0 (7.4) | +5.7 (4.2) | 318 | +5.5 (4.8) |
| L | Equilibrium | L3 | 226 | −88.1 (8.7) | −82.7 (7.5) | +5.4 (4.3) | 234 | +5.2 (4.5) |
| P | Venous | L1 | 97 | −85.9 (7.4) | −82.6 (6.8) | +3.3 (4.4) | 98 | +3.2 (4.4) |
| P | Venous | L3 | 96 | −87.8 (8.1) | −83.7 (7.0) | +4.1 (4.7) | 97 | +4.1 (4.7) |
| P | Equilibrium | L1 | 23 | −87.6 (6.3) | −81.5 (5.6) | +6.1 (2.6) | 23 | +6.1 (2.6) |
| P | Equilibrium | L3 | 22 | −88.7 (7.8) | −82.8 (7.0) | +5.9 (3.0) | 22 | +5.9 (3.0) |

#### *SAT attenuation (HU)*

| Dataset | Phase | Level | Patients | Non-contrast, mean (SD) | Contrast-enhanced, mean (SD) | Difference, mean (SD) | All examinations: pairs | All examinations: difference, mean (SD) |
|---|---|---|---|---|---|---|---|---|
| L | Early arterial | L1 | 308 | −88.7 (10.6) | −87.7 (10.5) | +1.0 (1.5) | 321 | +1.0 (1.6) |
| L | Early arterial | L3 | 229 | −91.7 (11.6) | −90.6 (11.5) | +1.2 (1.8) | 239 | +1.1 (2.0) |
| L | Late arterial | L1 | 309 | −88.7 (10.6) | −85.6 (10.4) | +3.1 (2.4) | 322 | +3.1 (2.7) |
| L | Late arterial | L3 | 228 | −91.7 (11.6) | −88.2 (11.8) | +3.5 (3.2) | 236 | +3.3 (3.4) |
| L | Portal venous | L1 | 310 | −88.7 (10.5) | −84.9 (10.4) | +3.8 (2.8) | 324 | +3.7 (2.9) |
| L | Portal venous | L3 | 311 | −92.1 (11.4) | −88.0 (11.4) | +4.1 (3.7) | 325 | +4.0 (4.0) |

| Dataset | Phase | Level | Patients | Non-contrast, mean (SD) | Contrast-enhanced, mean (SD) | Difference, mean (SD) | All examinations: pairs | All examinations: difference, mean (SD) |
|---|---|---|---|---|---|---|---|---|
| L | Equilibrium | L1 | 306 | −88.7 (10.6) | −84.1 (10.3) | +4.6 (3.5) | 318 | +4.5 (3.8) |
| L | Equilibrium | L3 | 226 | −91.7 (11.5) | −86.2 (11.5) | +5.5 (4.4) | 234 | +5.3 (4.9) |
| P | Venous | L1 | 97 | −86.5 (11.3) | −83.0 (11.1) | +3.5 (3.6) | 98 | +3.4 (3.6) |
| P | Venous | L3 | 96 | −90.1 (12.6) | −85.3 (12.4) | +4.7 (4.0) | 97 | +4.7 (3.9) |
| P | Equilibrium | L1 | 23 | −88.8 (10.5) | −82.7 (10.8) | +6.1 (2.4) | 23 | +6.1 (2.4) |
| P | Equilibrium | L3 | 22 | −93.9 (9.6) | −86.6 (11.3) | +7.3 (3.3) | 22 | +7.3 (3.3) |

All patients of Datasets L and P, irrespective of the training, validation and test splits. For each patient, phase and level, the earliest examination with measurements in both the non-contrast and the contrast-enhanced series was used. Values are fully automated (no manual correction) and as measured (no phase-0 conversion); the reference phase was used. The difference is the within-patient difference (contrast − non-contrast); 95% limits of agreement can be computed as mean ± 1.96 SD. Dataset L: 6 series without a recorded phase were excluded; per phase, 334–336 examinations had a contrast-enhanced series, 8 of which lacked a non-contrast series. At L1, 4–8 examinations lacked a measurement in one of the series. At L3, 88–92 examinations of the early arterial, late arterial and equilibrium phases lacked a measurement, mainly because these phases covered only the upper abdomen and L3 was outside the scan; in the portal venous phase, 3 did. An examination counted once if both series lacked a measurement. Dataset P: 1 examination lacked a non-contrast series, and at L3, 1 examination per phase lacked a measurement. Using all examinations added 8–14 pairs in Dataset L (later examinations of the same patients) and 0–1 in Dataset P; mean differences changed by ≤ 0.2 HU.

Protocol (both datasets): tube voltage 120 kVp; iodinated contrast medium at 600 mgI/kg (maximum 100 mL), injected over 30 s. Scan timing after the start of injection: Dataset L, early arterial phase by bolus tracking (about 25 s), late arterial phase 15 s later (about 40 s), portal venous phase 70 s and equilibrium phase 180 s; Dataset P, venous phase 90 s and, when acquired, equilibrium phase 180 s (the arterial phase of Dataset P was acquired in expiration and was not used).

These values differ in population from Table 3 and Fig. 6 of the main text, which used the test splits, all examinations and Datasets L, O and P.

### Table S16. Accuracy of the phase-0 conversion of areas

Median absolute percentage error (%) of the area against the non-contrast series of the same examination, as measured and after conversion, in the same-examination pairs of the test splits (datasets as in Table S11). For this evaluation, the power equations (Table S11) were refitted on the pairs of the training and validation splits only. Last two columns: median signed percentage error (positive, overestimation) of VAT area in the pairs in the lowest quartile of non-contrast VAT area, for each level and phase.

| Level | Phase | Test pairs | SMA: as measured | SMA: converted | VAT: as measured | VAT: converted | SAT: as measured | SAT: converted | VAT, lowest quartile: as measured | VAT, lowest quartile: converted |
|---|---|---|---|---|---|---|---|---|---|---|
| L1 | 1 Early arterial | 54 | 2.4% | 2.1% | 9.8% | 6.5% | 2.2% | 2.2% | −18.8% | +1.2% |

| Level | Phase | Test pairs | SMA: as measured | SMA: converted | VAT: as measured | VAT: converted | SAT: as measured | SAT: converted | VAT, lowest quartile: as measured | VAT, lowest quartile: converted |
|---|---|---|---|---|---|---|---|---|---|---|
| L1 | 2 Late arterial | 54 | 2.3% | 2.3% | 20.1% | 7.4% | 3.0% | 2.2% | −47.3% | +2.4% |
| L1 | 3 Portal venous | 50 | 2.8% | 2.6% | 21.0% | 7.7% | 3.6% | 3.3% | −41.4% | +27.0% |
| L1 | 4 Venous | 24 | 2.8% | 3.3% | 28.8% | 11.3% | 5.3% | 4.7% | −49.6% | +14.7% |
| L1 | 5 Equilibrium | 51 | 2.7% | 2.1% | 29.6% | 8.4% | 5.6% | 4.0% | −42.0% | +34.3% |
| L3 | 1 Early arterial | 49 | 1.8% | 2.1% | 11.0% | 3.8% | 1.4% | 1.8% | −20.1% | +1.4% |
| L3 | 2 Late arterial | 49 | 3.4% | 3.1% | 22.8% | 8.7% | 2.4% | 1.9% | −37.6% | +3.6% |
| L3 | 3 Portal venous | 50 | 2.2% | 2.0% | 21.9% | 6.6% | 2.7% | 2.9% | −41.3% | +14.5% |
| L3 | 4 Venous | 24 | 2.2% | 2.8% | 21.4% | 11.1% | 3.5% | 1.9% | −39.5% | +21.8% |
| L3 | 5 Equilibrium | 46 | 3.8% | 2.5% | 32.0% | 10.9% | 4.2% | 5.2% | −41.3% | +15.8% |

Percentage error, (value − non-contrast value) / non-contrast value × 100. Phase 0 (non-contrast) needs no conversion.

---

### Methods S4. Muscle and marrow age

**Rationale.** Chronological age is always known, so the information in an image-based age estimate lies in its difference from chronological age, which reflects the ageing of a particular tissue or organ. Deep-learning age estimation has been applied to images of many body regions [S1], and organ systems age at different rates. Organ-specific age gaps estimated from physiological and plasma protein data are associated with chronic disease and mortality [S2, S3], and age estimated from chest radiographs predicted mortality better than chronological age [S4]. On abdominal CT, a tissue age derived from muscle, fat and bone measures was associated with chronic conditions and mortality [S5].

**Models.** Four regression models (marrow and muscle at L1 and L3; Gaussian output; Table S1) were trained on Datasets L and main with ±20 HU intensity augmentation to reduce the effect of contrast, and combined by inverse-variance weighting, with the width of the 95% prediction interval calibrated on validation data. The models used in the prognostic analysis were trained without Dataset main (Table S1, note).

**Evaluation.** Mean absolute error (MAE), correlation, coverage of the 95% prediction interval, error by age decade and the effect of contrast (same examination, different phases in Dataset L) were assessed on the test splits of Datasets L and main, separately and pooled.

**Results.** The combined estimate had an MAE of 6.91 years in Dataset L (n = 253; r = 0.79; coverage of the 95% interval 0.98), 6.20 years in Dataset main (n = 301; r = 0.73; coverage 0.97) and 6.53 years pooled (n = 554; r = 0.76; coverage 0.98); the MAE of the muscle-only and marrow-only combinations was 7.05 years each (Fig. S4). Estimates regressed toward the mean, overestimating the age of patients in their twenties and underestimating that of patients in their eighties (Fig. S4a). Within the same examination, contrast-enhanced series were estimated 0.5–2.4 years younger than the non-contrast series (within-examination SD 1.44 years; 52 examinations; Fig. S4b). Muscle and marrow age gaps were moderately correlated (r = 0.44) and did not add to the prognostic model with low muscle mass and MSMD (HR per 5 years for OS 1.03 [95% CI 0.95–1.12] and 1.00 [0.94–1.06]; Tables S7 and S8).

**Interpretation.** The MAE was comparable to deep-learning age estimation from thoracic and abdominal CT (5.8–6.5 years) [S8]. The regression toward the mean requires that age gaps be defined as residuals [S6, S7], and contrast lowered the estimates by up to 2.4 years despite intensity augmentation. Age has also been estimated from vertebrae on CT for forensic purposes [S9]. To our knowledge, separate muscle and marrow age estimates have not been evaluated as prognostic markers; their value may lie in other settings, such as frailty or non-cancer outcomes.

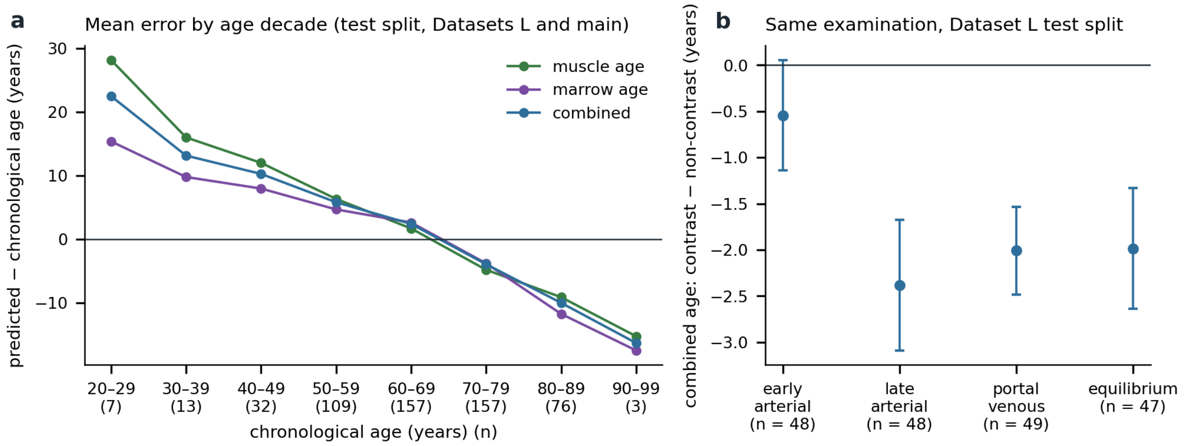


**Fig. S4** Muscle and marrow age. Mean error of the estimated age by decade of chronological age on the test splits of Datasets L and main (a), and the difference of the combined estimate between contrast-enhanced and non-contrast series of the same examination in Dataset L (b; mean and 95% CI).

### Methods S5. Comparison with TotalSegmentator

**Methods.** TotalSegmentator 2.18.0 [6, 26] was used under the TotalSegmentator Non-Commercial License for comparison only. The L3 vertebra was taken from the `total` task (1.5-mm model) and skeletal muscle from the `tissue_types` task. We compared (a) muscle on the reference slice and (b) each tool's own slice, which for TotalSegmentator was the craniocaudal middle of its L3 mask. The truncated test volumes were also processed with TotalSegmentator. Contrast-phase estimation was compared with `totalseg_get_phase` on the test splits of Datasets L, HC, O and P (422 series with a reference phase; 409 assessed by both methods). Series stored feet first were given to TotalSegmentator in head-first order. The proportional bias was assessed by regressing the difference on the mean (ordinary least squares).

**Agreement (Table S13, Fig. S3).** TotalSegmentator's muscle had a Dice of 0.846 ± 0.040 (0.888 ± 0.030 after thresholding) on the reference L3 slice and underestimated SMA by 10.7 cm$^2$ (9.9% of the reference mean; LoA −23.6 to 2.1 cm$^2$; ICC(A,1) 0.878; ICC(C,1) 0.964), with |ΔSMA| > 10 cm$^2$ in 1104 of 2009 series. The underestimation increased with muscle area (slope of the difference on the mean −0.084 [95% CI −0.096 to −0.073]; about −8 cm$^2$ at the 10th and −13 cm$^2$ at the 90th percentile); sarco showed no such trend (slope −0.000 [−0.007 to 0.007]). With each tool's own slice, in the 1989 series in which both tools found L3, the ICC(A,1) for SMA was 0.984 (sarco) and 0.879 (TotalSegmentator); TotalSegmentator's slice was within 5 mm of the reference slice in 47.0% of series (sarco, 95.3%), largely reflecting the different slice definitions.

**Phase estimation.** Four-group accuracy was 0.949 for sarco and 0.795 for TotalSegmentator (0.918 when the equilibrium phase, which TotalSegmentator does not output, was excluded). Sarco was more accurate for the arterial phases (0.949 vs 0.752) and TotalSegmentator for the portal venous and venous phases (1.000 vs 0.859); both separated non-contrast from contrast-enhanced series almost perfectly (0.998 and 1.000, binary accuracy). Accuracy by scan coverage and dataset is given in Table S4.

**Interpretation.** The underestimation agrees in direction with a multi-tool comparison in which TotalSegmentator consistently gave smaller SMA, with median absolute differences of about 15–21% from another automated tool [31], and TotalSegmentator's muscle Dice on our reference slices (0.846) was similar to the 0.83 reported on an external dataset [S10]. The high ICC(C,1) indicates that TotalSegmentator ranks patients similarly, but its absolute values, and thus thresholds such as that of Fujiwara et al. [23], are not interchangeable with values from corrected manual segmentation. Differences in slice selection were mostly due to definitions: TotalSegmentator's vertebral midpoint differs from the slice showing both transverse processes used in many clinical studies. Because our reference labels originate from an earlier version of the sarco model family, these comparisons may favour sarco.

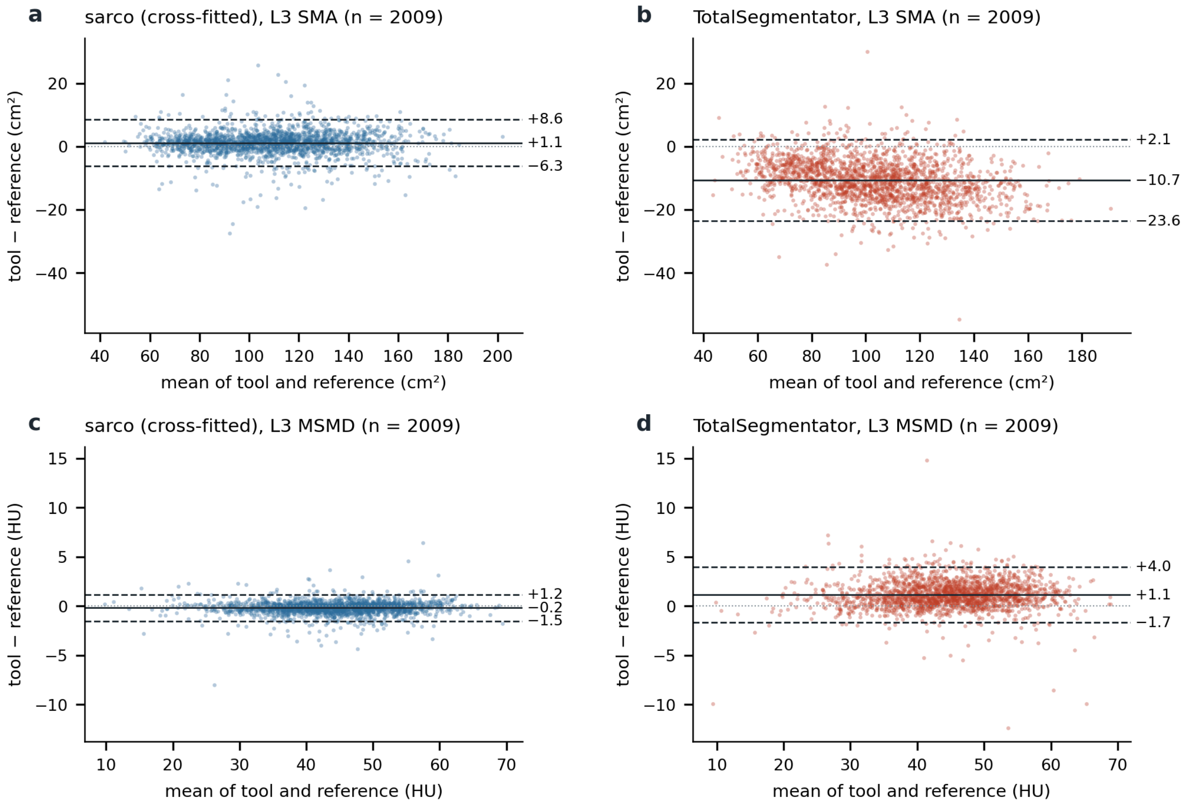


**Fig. S3** Bland–Altman plots on the reference L3 slice of Dataset main for SMA (a, b) and MSMD (c, d): sarco with cross-fitted models (a, c) and TotalSegmentator (b, d) against the reference labels (n = 2009). Solid lines, mean difference; dashed lines, 95% limits of agreement.

### Table S13. Agreement of TotalSegmentator with the reference labels (Dataset main, L3)

| | sarco (cross-fitted) | TotalSegmentator |
|---|---|---|
| (a) Reference slice: n | 2010 | 2009 |
| (a) Dice, muscle (thresholded) | 0.944 (0.964) | 0.846 (0.888) |

| | sarco (cross-fitted) | TotalSegmentator |
|---|---|---|
| (a) SMA ICC(A,1); ICC(C,1); bias [LoA], $cm^2$ | 0.988; 0.989; +1.15 [−6.27, 8.57] | 0.878; 0.964; −10.7 [−23.6, 2.1] |
| (a) Slope of the SMA difference on the mean [95% CI] | −0.000 [−0.007, 0.007] | −0.084 [−0.096, −0.073] |
| (b) Own slice: n | 2007 | 1990 |
| (b) SMA ICC(A,1); bias [LoA], $cm^2$ | 0.983; +1.35 [−7.44, 10.14] | 0.879; −8.1 [−27.2, 11.0] |
| (b) Slope of the SMA difference on the mean [95% CI] | −0.001 [−0.009, 0.006] | −0.016 [−0.033, 0.001] |
| (b) Slice within 5 mm of the reference | 1912/2007 (95.3%) | 936/1990 (47.0%) |

TotalSegmentator: 2009 series with output (one failure); 1990 with L3 found by TotalSegmentator; 1989 with L3 found by both tools (SMA ICC(A,1) with own slices in these 1989 series: sarco 0.984, TotalSegmentator 0.879). LoA, 95% limits of agreement.

### References S1–S10


S1. Haugg F, Lee G, He J, Johnson J, Zapaishchykova A, Bitterman DS, et al. Imaging biomarkers of ageing: a review of artificial intelligence-based approaches for age estimation. Lancet Healthy Longev. 2025;6:100728. https://doi.org/10.1016/j.lanhl.2025.100728

S2. Tian YE, Cropley V, Maier AB, Lautenschlager NT, Breakspear M, Zalesky A. Heterogeneous aging across multiple organ systems and prediction of chronic disease and mortality. Nat Med. 2023;29:1221–31. https://doi.org/10.1038/s41591-023-02296-6

S3. Oh HS, Rutledge J, Nachun D, Pálovics R, Abiose O, Moran-Losada P, et al. Organ aging signatures in the plasma proteome track health and disease. Nature. 2023;624:164–72. https://doi.org/10.1038/s41586-023-06802-1

S4. Raghu VK, Weiss J, Hoffmann U, Aerts HJWL, Lu MT. Deep learning to estimate biological age from chest radiographs. JACC Cardiovasc Imaging. 2021;14:2226–36. https://doi.org/10.1016/j.jcmg.2021.01.008

S5. Rule AD, Grossardt BR, Weston AD, Garner HW, Kline TL, Chamberlain AM, et al. Older tissue age derived from abdominal computed tomography biomarkers of muscle, fat, and bone is associated with chronic conditions and higher mortality. Mayo Clin Proc. 2024;99:878–90. https://doi.org/10.1016/j.mayocp.2023.09.021

S6. Le TT, Kuplicki RT, McKinney BA, Yeh HW, Thompson WK, Paulus MP, et al. A nonlinear simulation framework supports adjusting for age when analyzing BrainAGE. Front Aging Neurosci. 2018;10:317. https://doi.org/10.3389/fnagi.2018.00317

S7. Smith SM, Vidaurre D, Alfaro-Almagro F, Nichols TE, Miller KL. Estimation of brain age delta from brain imaging. Neuroimage. 2019;200:528–39. https://doi.org/10.1016/j.neuroimage.2019.06.017

S8. Kerber B, Hepp T, Küstner T, Gatidis S. Deep learning-based age estimation from clinical computed tomography image data of the thorax and abdomen in the adult population. PLoS One. 2023;18:e0292993. https://doi.org/10.1371/journal.pone.0292993

S9. Kawashita I, Fukumoto W, Mitani H, Narita K, Chosa K, Nakamura Y, et al. Development of a deep-learning algorithm for age estimation on CT images of the vertebral column. Leg Med (Tokyo). 2024;69:102444. https://doi.org/10.1016/j.legalmed.2024.102444

S10. Chen Y, Gu H, Chen Y, Yang J, Dong H, Cao JY, et al. Automated muscle and fat segmentation in computed tomography for comprehensive body composition analysis. Mach Learn Biomed Imaging. 2025;2025:581–618. https://doi.org/10.59275/j.melba.2025-ag5g
